# Impact of non-pharmaceutical interventions on COVID-19 incidence and deaths: cross-national natural experiment in 32 European countries

**DOI:** 10.1101/2022.07.11.22277491

**Authors:** Diogo Costa, Sven Rohleder, med Kayvan Bozorgmehr

## Abstract

**Purpose:** Non-pharmaceutical interventions (NPIs) have been the cornerstone of COVID-19 pandemic control, but evidence on their effectiveness varies according to the methods and approaches taken to empirical analysis.

We analysed the impact of NPIs on incident SARS-CoV-2 across 32 European countries (March-December 2020) using two NPI trackers: the Corona Virus Pandemic Policy Monitor – COV-PPM, and the Oxford Covid-19 Government Response Tracker – OxCGRT.

**Methods:** NPIs were summarized through principal component analysis into three sets, stratified by two waves (C1-C3, weeks 5-25, and C4-C6, weeks 35-52). Longitudinal, multi-level mixed-effects negative binomial regression models were fitted to estimate incidence rate ratios for cases and deaths considering different time-lags and reverse causation (i.e. changing incidence causing NPIs), stratified by waves and geographical regions (Western, Eastern, Northern, Southern, Others).

**Results:** During the first wave, restrictions on movement/mobility, public transport, public events, and public spaces (C1) and healthcare system improvements, border closures and restrictions to public institutions (C2) reduced SARS-CoV-2 incidence after 28 and 35-days. Mask policies (C3) reduced SARS-CoV-2 incidence (except after 35-days). During wave 1, C1 and C2 reduced deaths after 49-days and C3 after 21, 28 and 35-days. During wave 2, restrictions on movement/mobility, public transport and healthcare system improvements (C5) decreased SARS-CoV-2 cases and deaths across all countries.

**Conclusion:** In the absence of pre-existing immunity, vaccines or treatment options, the impact of NPIs on SARS-CoV-2 incidence and deaths varied by regions and waves but was consistent across components of NPIs derived from two policy trackers (CoV-PPM and OxCGRT).

## Introduction

The COVID-19 pandemic has triggered a broad range of non□pharmaceutical interventions (NPI), i.e. population-level policies and measures, that aim to prevent and/or control SARS-CoV-2 transmission among individuals and communities.[1] As the pandemic unfolded, more evidence from observational studies emerged, assessing the relative importance and contribution of measures simultaneously implemented.

A systematic review of the methodologies used to assess the effectiveness of NPIs during COVID-19 identified a scarcity of subgroup analysis, necessary to depict differences in effectiveness variation, and also calls for studies where variation in methodologies can be applied, namely through sensitivity analysis that repeat the same analysis with different sets of publicly available NPIs datasets[2]. Other methodological shortcomings challenged by the quality of the outcome data used, which could have been influenced by changes in (surveillance) systems capacities, changing rules or delayed reporting, and that could be at least partly tested by the use of different outcomes within the same study, or testing alternate lag periods for the impact of NPIs [3]. This may help to identify the influence of certain methodological approaches and strengthen the evidence in favour of the effectiveness of particular NPIs.

Previous studies that included European countries found that physical distancing was associated with a reduction in COVID-19 incidence [4]. Restrictions on gatherings, closing of specific sectors (e.g., restaurants, schools, kindergartens, etc) and closing of some or all school levels were also found to reduce the epidemic growth rare across the 37 Organisation for Economic Co-operation and Development (OECD) members, in an early phase (October-December 2020) [5]. Another study analysing NPI effectiveness across 30 European countries found that a combination of measures involving school closures, banning mass gatherings and early closure of commercial businesses was associated with reduced infections, but other measures, like extensive closure of all non-essential business and stay-at-home orders, were not [6]. These efforts have been confined to the analysis of single outcomes, of single specific measures, single countries, or specific regions, so there is still need to explore the type, combination, or degree of implementation of NPIs that has been effective to mitigate the transmission of SARS-Cov-2 or associated deaths at population level throughout the infection waves.

Most data have been made publicly available during the COVID-19 pandemic, which allows testing different methodologies, conducting sensitivity analysis to the effectiveness of the same NPIs, even if differently framed, categorised, and recorded, and seek for commonalities within subgroups of the population (for example, groups of neighbouring, culturally close countries, sharing borders, climate and other contextual features with potential pandemic impact). Outcome data availability also has its shortcomings, since the quality of reporting and recording systems is not assured and bound to changes. Hence, subgroup testing, consideration for varying timing of effects and analytical approaches aiming to rule-out the potential for reverse causation are needed.

In this study, we contribute to the rapidly growing field of evidence on NPI effectiveness, taking some of these methodological concerns into account. We exploit the heterogeneity in timing, temporal sequence, and combination of measures within and across 32 European countries as a natural experiment to assess which combinations of NPIs have been effective to reduce SARS-Cov-2 incidence and associated deaths at the population-level in the early phases of the pandemic. Furthermore, we investigate subgroups of countries (according to geographic region) and apply the same analysis to two different NPIs data sources.

## Methods

### Study design

The study design resembles a natural experiment[7], in which the populations in countries were repeatedly exposed to different timing and combination of NPIs which ultimately pursued a common aim: reducing population-level transmissions of SARS-CoV-2 and related deaths. Therefore, the effects on outcomes can be studied using each country as their own control, and each country for controls of other countries with different timing or combination of exposures.

### Data sources

We resorted to the Corona Virus Pandemic Policy Monitor (COV-PPM) that prospectively monitors and tracks NPIs in 32 countries of the EU27, EEA and UK [8]. A total of eight NPI categories (Panel 1) were retrieved until December 2020, with different sub-categories covering relevant areas of societal living. For each NPI category exact starting dates and duration of implementation are registered in a daily format. A detailed description of methods, data validation process, and usage options can be found elsewhere [8]. The subcategories of retrieved NPIs categories (Panel 1) were combined into categorical variables for analysis as shown in supplementary material (Panel S1).

##### Panel 1: Categories of non-pharmaceutical interventions (NPI)

a) Restrictions to public events, namely in the number of persons allowed for indoor/outdoor events (conferences, sports, festivals), for more than 1000 persons, less than 1000 persons, more than 50 persons, for any number or without specification;
b) Restrictions or closures to public institutions (incl. schools, universities, public services), in single cities, at the state level, nationally or without specification;
c) Restrictions and closures in public spaces (incl. shops, bars, gyms, restaurants), in single cities, at the state level, nationally or without specification;
d) Measures affecting public transport (incl. trains, buses, trams, metro), in single cities, at the state level, nationally or without specification;
e) Restrictions in movement/mobility, to pedestrians, private cars, national aviation, other means or without specification;
f) Border closures or restrictions applied to travelling by air, land or sea, across national borders, for non-nationals from high-risk regions, for all non-nationals, for all incoming travellers or without specification;
g) Measures relating to Human Resources reinforcement in healthcare (incl. human resources reinforcement or redistribution, technical reinforcement or redistribution, material infrastructural reinforcement or without specification);
h) Masks (mandatory or recommended use of facial and nose protection);

To triangulate measures obtained from other NPI trackers (and allow a posterior sensitivity analysis of models using two distinct exposure measures), we also resorted to the Oxford Covid-19 Government Response Tracker (OxCGRT)[10], and used the specific categories “C1-School closing”, “C2-Workplace closing”, “C3-Cancel public events”, “C4-Restrictions on gatherings”, “C5-Close public transports”, “C7-Restrictions on internal movement”, “C8-International travel controls” and “H6-Facial coverings” (version downloaded on 31.05.2021).

A total of 8512 country-days in 32 countries (see supplementary material Figures S1-8) were analysed stratified by two periods of pandemic waves during 2020 (wave 1: from calendar week 5 to 25, i.e. end of February to June; wave 2: week 35 to 52, i.e. end of August to December). The daily number of notified SARS-Cov-2 cases and associated deaths in each country, were retrieved from the WHO [9]. A 7-day smoothed average of reported cases was used as main outcome, to accommodate the expected week-weekend variation in case notification. Country population size (2019) and selected macroeconomic indicators were retrieved from EUROSTAT (https://ec.europa.eu/eurostat/web/main/data/database) for the most recent period available: Gross domestic product (GDP) per capita (Current market prices, million euro, 2019), Health care expenditure (Million Euro per habitant 2017), and Population density (inhabitants per square kilometre, 2018).

### Principal component analysis

A principal component analysis (PCA) was conducted to reduce the data collected in the scope of COV-PPM (categories shown in Panel 1) into relevant related factors, reflecting periods when distinct NPIs were simultaneously implemented. The scree plots and percentage of variance explained were analysed for each wave to decide the number of components to extract (supplementary material Figure S9). Orthogonal varimax rotation was conducted to identify individual NPIs loadings in each component. Each component score was then scaled by 10 and conversed into scores with non-negative values and a mean of 50.

A descriptive analysis of (means, standard deviations, and within/between country variation over time of NPI categories and derived component scores obtained with COV-PPM are shown in supplementary material, Table S1).

The OxCGRT categories of NPIs “C1-School closing”, “C2-Workplace closing”, “C3-Cancel public events”, “C4-Restrictions on gatherings”, “C5-Close public transports”, “C7-Restrictions on internal movement”, “C8-International travel controls” and “H6-Facial coverings” were entered in a PCA analysis for data reduction, following the same procedure implemented for COV-PPM data. Tables 1 and 2 show the scoring coefficients of the resulting components obtained using each NPIs database, respectively.

**Table 1.** Scoring coefficients for orthogonal varimax rotation, non-pharmaceutical interventions (Corona Virus Pandemic Policy Monitor - COV-PPM)

| Variable | Wave 1 |  |  | Wave 2 |  |  |
| --- | --- | --- | --- | --- | --- | --- |
|  | Component 1 | Component 2 | Component 3 | Component 4 | Component 5 | Component 6 |
| Public events | 0.3660 | 0.1656 | 0.1253 | 0.5822 | -0.1659 | 0.0838 |
| Public institutions | 0.2989 | 0.3208 | -0.0227 | 0.3271 | 0.2379 | 0.1034 |
| Public spaces | 0.3616 | 0.2070 | 0.1225 | 0.5327 | -0.0887 | 0.1383 |
| Public transport | 0.4371 | 0.0344 | 0.0079 | 0.1726 | 0.4868 | 0.0261 |
| Movement/mobility | 0.6512 | -0.2768 | -0.1401 | -0.1384 | 0.7520 | 0.0805 |
| Border closure/Travelling | 0.0899 | 0.4549 | 0.1454 | 0.4218 | 0.0453 | -0.2899 |
| Healthcare system improvement | -0.1475 | 0.7349 | -0.1385 | 0.1995 | 0.3126 | -0.3943 |
| Masks | -0.0305 | -0.0443 | 0.9533 | 0.0606 | 0.0765 | 0.8465 |

**Table 2.** Scoring coefficients for orthogonal varimax rotation, non-pharmaceutical interventions (Oxford

Covid-19 Government Response Tracker - OxCGRT)
| Variable | Wave 1 |  |  | Wave 2 |  |  |
| --- | --- | --- | --- | --- | --- | --- |
|  | Component 1-Ox | Component 2-Ox | Component 3-Ox | Component 4-Ox | Component 5-Ox | Component 6-Ox |
| School closings | 0.3827 | 0.1556 | 0.0308 | 0.2754 | -0.4226 | 0.0660 |
| Workplace closing | 0.3531 | 0.2041 | -0.0144 | 0.4335 | 0.0738 | 0.0609 |
| Cancel public events | 0.4741 | -0.0304 | 0.0566 | 0.4155 | 0.2938 | 0.1879 |
| Restrictions in gatherings | 0.3601 | 0.1686 | 0.0718 | 0.4697 | 0.2355 | 0.0026 |
| Public transport | -0.0856 | 0.7835 | -0.0693 | 0.1060 | -0.0906 | 0.7948 |
| Internal Movement | 0.1370 | 0.4520 | 0.0762 | 0.3678 | -0.3494 | 0.1022 |
| Int Travel control | 0.5901 | -0.2933 | -0.1088 | 0.0910 | 0.7167 | -0.0308 |
| Facial coverings | -0.0118 | -0.0248 | 0.9839 | 0.4339 | -0.1738 | -0.5600 |

Three components per wave, i.e. sets of combinations of NPIs, explained 74% of the variance for the first (C1-3) wave and 70% of the variation in the second wave (C4-6) for COV-PPM categories.

In the first wave, C1 was strongly related with NPIs referring to restrictions in movement/mobility (scoring coefficient for orthogonal varimax rotation: 0.6512), and, to a lesser extent, related with measures affecting public transport (0.4371), public events (0.3660) and public spaces (0.3616). C2 was mainly related with measures that aimed at improving the healthcare system (0.7349), border closure/travelling restrictions (0.4549) and measures impacting public institutions functioning (0.3208). C3 was related with recommendations or enforcement of mask utilization (0.9533).

In the second wave, C4 was related with restrictions in public events (0.5822), in public spaces (0.5327), border closures/travelling restrictions (0.4218) and public institutions (0.3271). C5, was related to restrictions to movement/mobility (0.7520), public transport (0.4868) and healthcare system improvement measures (0.3126). C6, was also primarily related with masks (0.8465).

For the OxCGRT, in wave 1, C1-Ox was related to measures on international travel control (0.5901), public events restrictions (0.4741), restrictions in gatherings (0.3601), school closing (0.3827) and workplace closing (0.3531). C2-Ox, was related to restrictions on public transport (0.7835) and internal movement (0.4520). C3-Ox was related to facial coverings (0.9839). In wave 2, C4-Ox was mainly related to restrictions in gatherings (0.4697), facial coverings (0.4339), workplace closing (0.4335), cancelation of public events (0.4155), and internal movement restrictions (0.3678). C5-Ox was primarily related to international travel control (0.7167) and C6-Ox was related to public transport related measures (0.7948).

### Statistical analysis

Scatter plots were used to explore the temporal change in NPIs as measured with COV-PPM and SARS-Cov-2 incidence (per 100,000) by country (supplementary material, Figures S1 – S8). For descriptive purposes, the proportion of observation time in which COV-PPM NPIs were in place, and the within and between country variation therein, was calculated by country and wave (supplementary material, Table S1).

A panel analysis was implemented to analyse the effect of NPIs on daily SARS-CoV-2 incidence and associated deaths by calculating incidence rate ratios (IRR) and corresponding 95% confidence intervals (CI). The IRR estimates hence quantify the relative difference in SARS-CoV-2 incidence or associated deaths for a one-unit change in component scores over time a) *between* countries, comparing countries with different NPI component scores, and b) *within* countries, comparing variation in respective NPIs implemented and their stringency (simultaneous presence of categories of each measure).

Different model specifications fitted to the data were tested (Supplementary material, Table S2 and description). A multi-level mixed-effects negative binomial model with country as random-intercept showed the best model fit according to lowest Akaike (AIC) and Bayesian Information Criteria (BIC) (supplementary material, Table S2). The analysis was guided by a causal diagram, illustrating the potential causal/non-causal pathways from NPIs implementation to outcomes over time (Figure 1). Crude models were fitted including the components obtained by PCA (“immediate” or baseline effects) and the outcome. These were further adjusted for one of five time-lagged variables of the same components in five separate models, respectively (crude model, and models adjusted for 7-, 14-, 21-, 28-, and 35-days lag of each component, respectively). The crude models (no lag) show the immediate association with the outcome of each component, adjusted for the effect of one another. The IRR estimates obtained from the further adjusted models show the independent lagged effect of each component, i.e. adjusted for the non-lagged association of each component with the outcome (incidence or deaths) and the lagged effect of the other components. The lags considered for deaths were 21-, 28-, 35-, 42-, and 49-days. The choice for 7-days increasing lagged-effect was based on SARS-Cov-2 natural history and the existing literature [11,12].

**Fig. 1.**
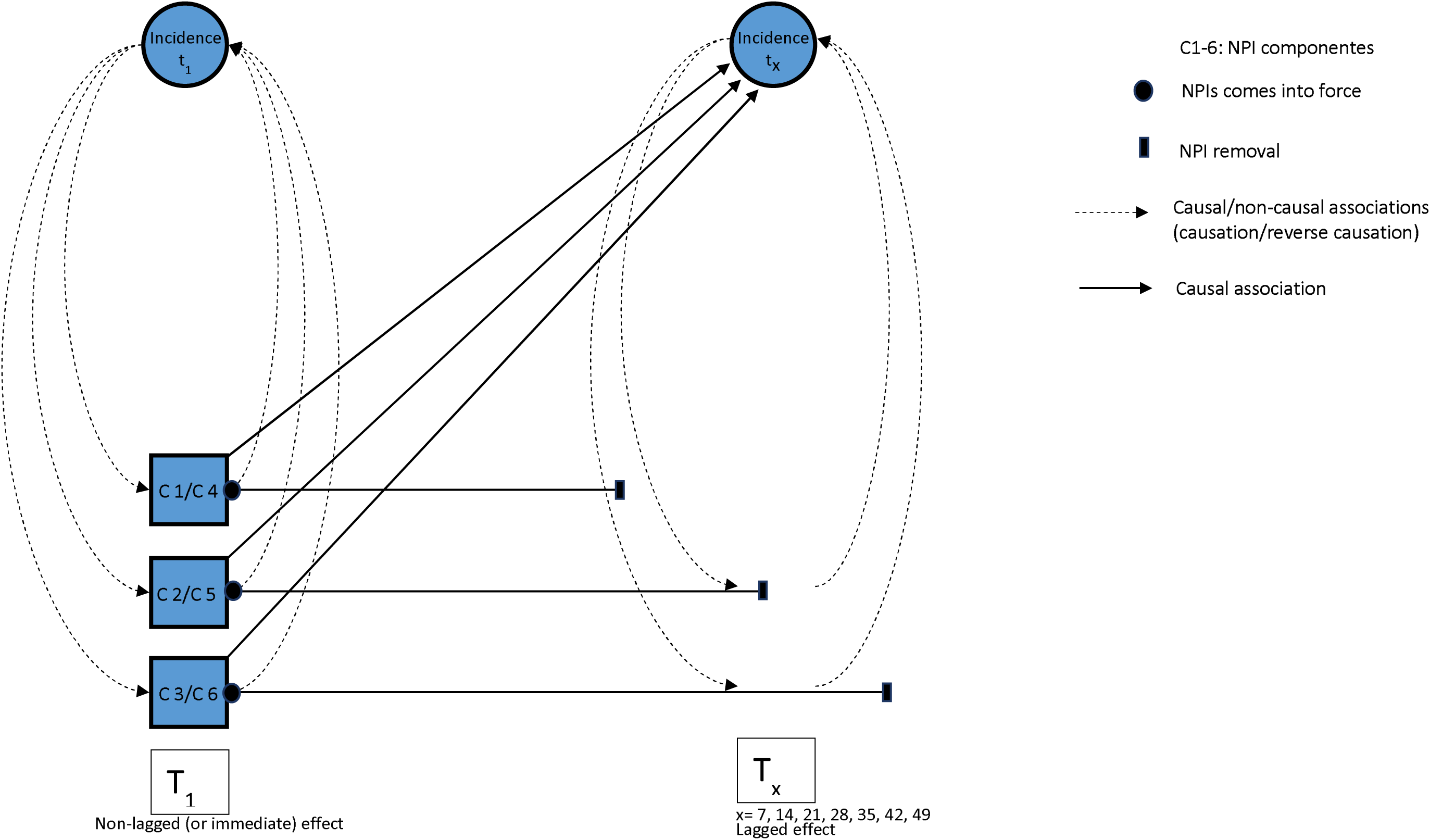
Causal diagram

All models further included time (in days) as discrete variable to consider secular trends by unobserved variables, the population size (as offset), and country (as random intercept). NPIs have often been implemented reactively in face of rising incidence, and have been lifted in view of declining incidence raising issues of reverse causation when studying their impact on infection dynamics. Therefore, a variable for the change rate in SARS-CoV-2 incidence 7 days before (see calculation below) was also entered in all models to account for potential reverse causation, i.e. preceding growth or decline in in incidence change rates impacting the introduction or removal of NPIs.

The mixed-effects negative binomial models with *Y_it_ ∼ NB(P_i_θ_it_)* where *Y_it_* are the observed Covid-19 cases or deaths, *θ_it_* the rate, and *P_i_* the country population size, were specified as follows.

For country *i* = {1,…, 32}, and day *t* = {1,…, 280} the rate *η_it_* = *log(θ_it_)* of observed cases or deaths *Y_it_*, and population size *P_i_* used as an offset log(*P_i_)* was specified on a logarithmic scale as:

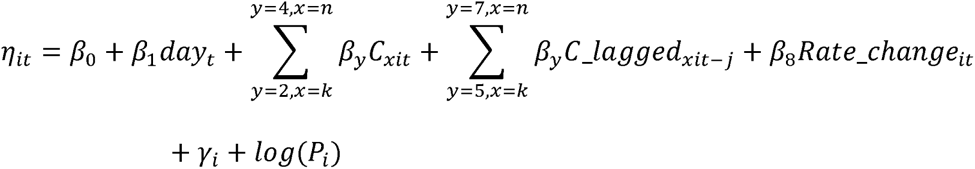

where *β*_0_ is the intercept, *β*_1_ is the coefficient of the identifier variable for days *day_t_*, *β*_2_ to *β*_4_ are the coefficients for the three component scores *C_xit_* of each wave, where component identifier *x* = {1, 2,3} used for modelling the first wave (i.e. *C_lit_*, *C_2it_* and *C_3it_*), and *x* = {4, 5,6} used for modelling the second wave (i.e. *C_4it_*, *C_Sit_* and *C_6it_*). *k* and *n* are the lower and upper limit of summation for the respective value set of *x*. *β*_5_ to *β*_7_ are the coefficients for the temporal lagged component scores *C_agged_xit-j_* using different component identifier *x* for the first and second wave (see above), while lagging the respective component score by j-days (for cases *j* = {7,14, 21,28, 35}, and for deaths *j* = {21,28,35,42,49}), which were entered in separate models for each level of *j*. *β*_8_ is the coefficient for the change in rate *Rate_change_it_* in Covid-19 incidence 7 days before, i.e. *Rate_change_it_* = *Y_it_*/*Y_it-7_* (entered in models for cases and deaths). *γ_i_* is the random effect modelled using exchangeability among countries, and *log(P_i_)* is the country population size entered as offset.

GDP, healthcare expenditure per capita, and population density were further tested as covariates, but not included in the final models since they did not improve model fit nor change the magnitude or direction of considered effects (supplementary Tables S12 and S13).

The final models were additionally stratified according to geographical regions in Europe: Southern (Portugal, Spain, Italy, Greece and Cyprus), Western (Belgium, Netherlands, France, Germany, Ireland, United Kingdom and Austria), Eastern (Czech Republic, Slovakia, Slovenia, Poland, Romania, Hungary and Bulgaria), Northern (Norway, Sweden, Finland and Denmark) and Other regions (Croatia, Estonia, Iceland, Latvia, Lithuania, Luxembourg, Malta, Switzerland and Liechtenstein).

All analysis were conducted using Stata 16®[13], visualisations were performed with R programming language 4.1.2. Geographic data for maps were retrieved from Eurostat [14].

## Results

### Descriptive analysis

Across all countries, a total of 1,614,594 COVID-19 cases and 178,369 associated deaths were analysed during the first wave and 18,471,042 cases and 328,426 deaths during the second wave (Figure 2). The timing of NPI implementation and the proportion of days during the observation period in which measures were in place across countries varied widely within and between countries during the two infection waves. In wave 1, restrictions to public events, public institutions, and public spaces were the most widely used measures (Figure 2, and supplementary material Table S1). In wave 2, these measures were partly relaxed, while mask policies, restrictions to travelling and border closures, and movement/mobility restriction became more prominent (Figure 2). Within and between country variation in NPIs remained high with the exception of masks where variation (especially within countries) was lower (supplementary material, Table S1).

**Fig. 2.**
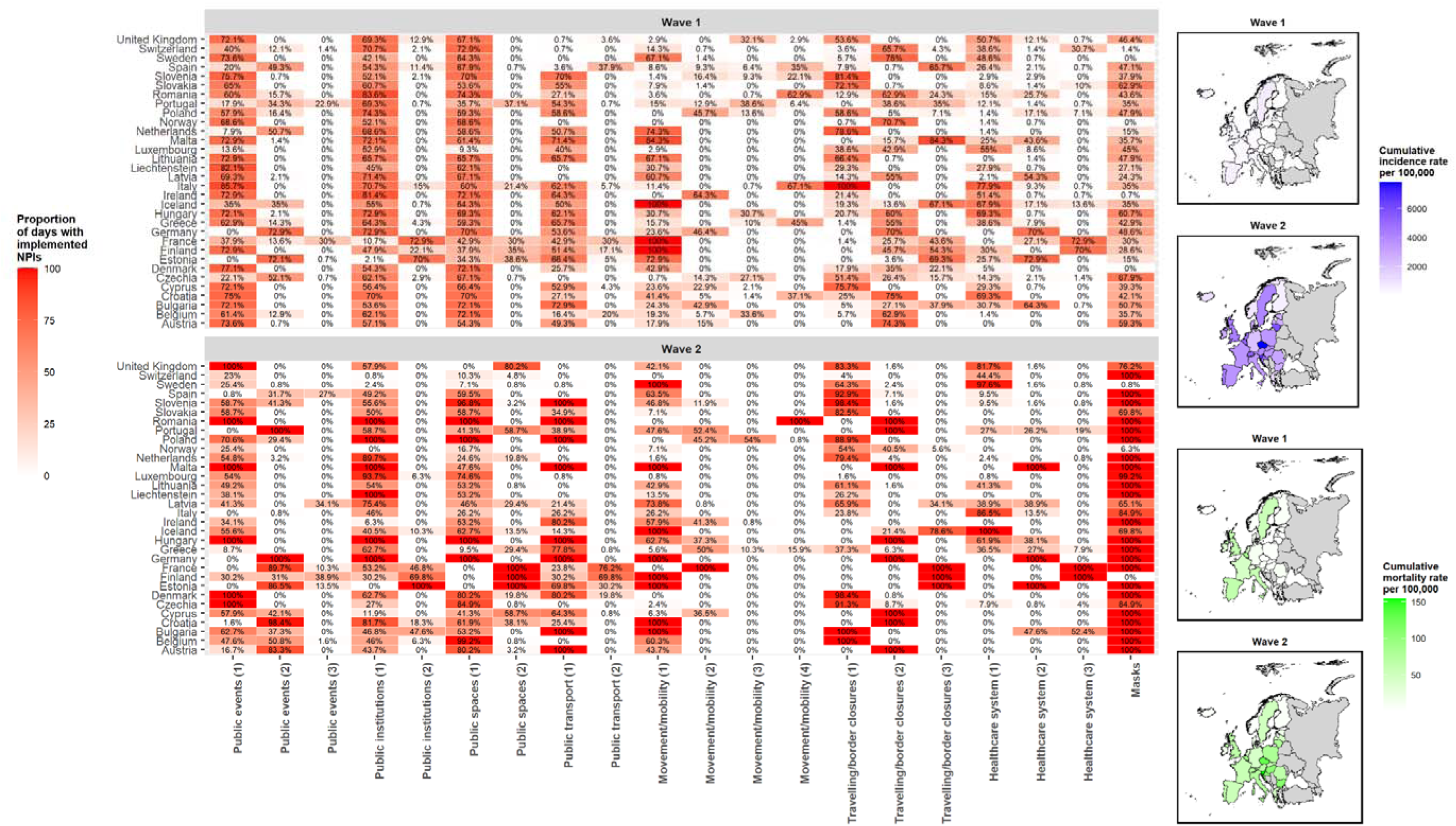
Proportion of days during the observation period in which NPIs were in place (Wave 1 - weeks 5-25, and Wave 2 – weeks 35-52), and cumulative incidence and mortality rates per 100,000 of SARS-Cov-2, in 32 countries (EU-27, EEA, UK) Legend: (1), (2), (3) and (4), refer to the simultaneous presence of sub-categories in place for each non-pharmaceutical intervention (i.e., 1: if at least one of the subcategories was present, 2: if at least two of the subcategories were simultaneously present, 3: if 3 subcategories were simultaneously present, and 4: if 4 subcategories were simultaneously present);

### NPI effects on SARS-CoV-2 Incidence

Incidence rate ratios (IRR) for the lagged-effects of the PCA-components of NPIs derived from COV-PPM, considering a 7, 14, 21, 28 and 35 days-lag across all countries and stratified by waves and regions are presented in Figure 3.

**Fig. 3.**
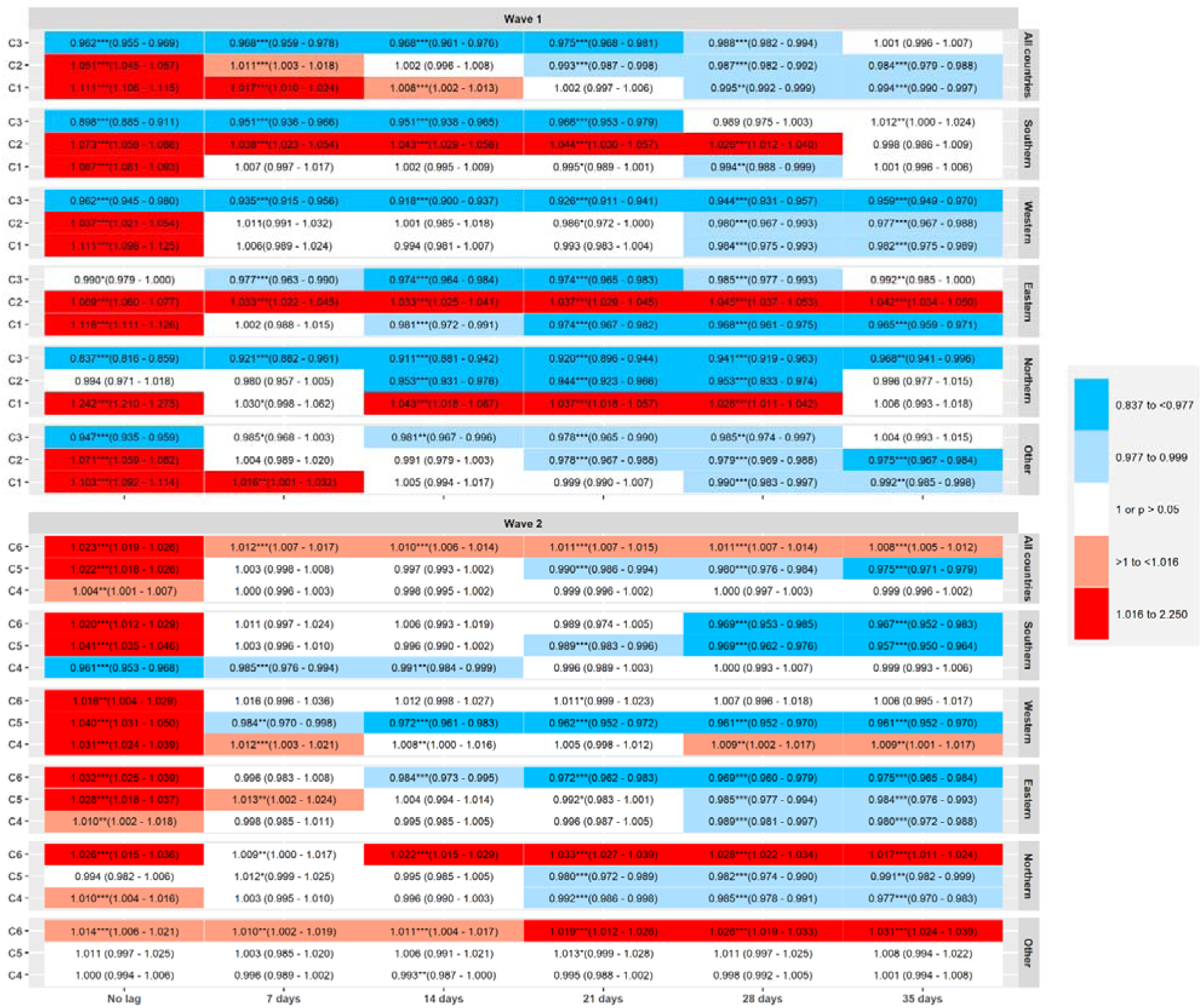
Incidence Rate Ratios (IRR) and 95% CI for the lagged-effects (7, 14, 21, 28 and 35 days) on SARS-CoV2 incidence of the three principal component (PCA) scores of NPIs, N=20,085,636 SARS-CoV-2 cases in 32 countries during the first and second waves of infections (March-December 2020), by region All countries: includes all countries of EU27, EEA and UK; Southern: Portugal, Spain, Italy, Greece and Cyprus; Western: Belgium, Netherlands, France, Germany, Ireland, United Kingdom and Austria; Eastern: Czech Republic, Slovakia, Slovenia, Poland, Romania, Hungary and Bulgaria; Northern: Norway, Sweden, Finland and Denmark; Other: Croatia, Estonia, Iceland, Latvia, Lithuania, Luxembourg, Malta, Switzerland and Liechtenstein. Within each regional stratum and column for time lags, IRR estimates represent the independent effects of C1-C3 (wave 1) and C4-C6 (wave 2), i.e. mutually adjusted for one another, and additionally adjusted for the immediate effect of each component (no lag), time, 7-day rate change in incidence, and country as random intercept.

Considering the findings across all countries during the first wave, the NPI combination C1 (movement/mobility, public transport, public events, public spaces) significantly reduced SARS-CoV-2 incidence in models considering a 28-days and 35-days lagged effect (adjusted for the baseline effect): IRR, 95%CI=0.995, 0.992-0.999 for the 28-day-lagged variable, 0.994, 0.990-0.997 for 35-day-lag). C2 (healthcare system improvement, border closures and restrictions in public institutions) revealed the same pattern as C1 across all countries, with significant association with lower incidence, IRR after 21-days (0.993, 0.987-0.998), 28-days (0.987, 0.982-0.992) and 35-days (0.984, 0.979-0.988).

C3 (masks) was associated with lower incidence in the non-lagged model (IRR=0.962, 0.955-0.969) and for all the time-lags considered except for a 35-days lag.

In southern countries during the first wave (Figure 3), C1 reduced incidence after 28-days, while C2 came along with higher incidence after 7, 14, 21 and 28 days-lag. C3 reduced SARS-CoV-2 incidence for all time-lags considered except after a 28 and 35-days lag.

In Western countries, C1 and C2 reduced incidence after 28 and 35 days, while C3 significantly reduced incidence for all time-lags considered.

In the Eastern region, C1 and C3 significantly reduced incidence rates for all lags considered (except C1 for “no-lag” effect), while C2 was associated with an increase in incidence for all lagged variables.

In Northern countries, C1 showed associations with an increase in incidence rates for 14, 21 and 28-days lag, while C2 and C3 significantly reduced incidence for all lags considered (except C2 after 7 days and after 35 days-lag, where no significant effect was observed).

In the remaining countries analysed as residual group (Croatia, Estonia, Iceland, Latvia, Lithuania, Luxembourg, Malta, Switzerland and Liechtenstein), C1 and C2 significantly reduced incidence after 28 and 35 days while C3 significantly reduced incidence after 14, 21 and 28 days.

Results of all full models estimates (i.e., including estimates for non-lagged effects for all models and corresponding 95% confidence intervals, CI) are shown in supplementary material (Tables S3, S4 for all countries and Table S8, S9, stratified by geographical regions and S12, S13 adjusted for GDP, healthcare expenditure and population density).

In the second wave (Figure 3), restrictions in public events, public spaces, border closures and restrictions in public institutions (C4) had no significant effect on incidence for any of the time-lags considered in all countries (except for the association with an increase in incidence in the non-lagged variable).

Restrictions in movement/mobility, public transport and healthcare system improvement measures (C5) significantly reduced incidence following 21-days (0.990, 0.986-0.994) 28-days (0.980, 0.976-0.984) and 35-days (0.975, 0.971-0.979). C6 (masks) unfolded significant associations with an increase in incidence for all time-lags considered, with the point estimate showing a decrease in magnitude with increasing lagged days.

In the Southern region, C4 significantly reduced incidence in the non-lagged model and after 7- and 14-days, but showed no effects for the remaining lags considered. C5 and C6 significantly reduced incidence after 21 (for C5), 28 and 35-days.

In the Western region, C4 increased incidence for all time-lags except 21-days. C5 significantly reduced incidence for all the time lags considered while C6 showed no significant effect on incidence for all the time lags considered.

In the Eastern region, the components C4-C6 showed a trend suggesting lower incidence according to the increase in time-lag considered, with significant reductions in incidence for C6 after 14, 21, 28 and 35 days and for C4 and C5 after 28 and 35 days. In the Northern region, C4 and C5 significantly reduced incidence after 21, 28 and 35 days, and C6 showed an association with increased incidence for all the time-lags considered.

In the “other” regions, C4 and C5 showed no effects on SARS-CoV-2 incidence (except for C4 after 14 days associated with lower incidence), while C6 showed significantly higher incidence for all time-lags analysed.

### Associated deaths

Considering all countries in the first wave (Figure 4), C1 was associated with an increase in deaths after a 21-,28-, and 35-days lagged effect (IRR, 95%CI for no lag=1.124, 1.119-1.129, for 21-days lag=1.037, 1.033-1.040, for 28-days=1.019, 1.015-1.022, for 35-days-lag=1.008, 1.004-1.011), which then turned into an association with a decrease in deaths after 49-days (0.994, 0.991-0.997). C2 showed the same trend as C1, associated with an increase in deaths for no-lag (1.060, 1.053-1.067), 21-days (1.014, 1.009-1.018), 28 days (1.007, 1.002-1.012) and with a decrease after 49-days (0.990, 0.986-0.995). C3 significantly reduced deaths for all the time lags considered across all countries, except after 42 and 49 days, when estimates were non-significant.

**Fig. 4.**
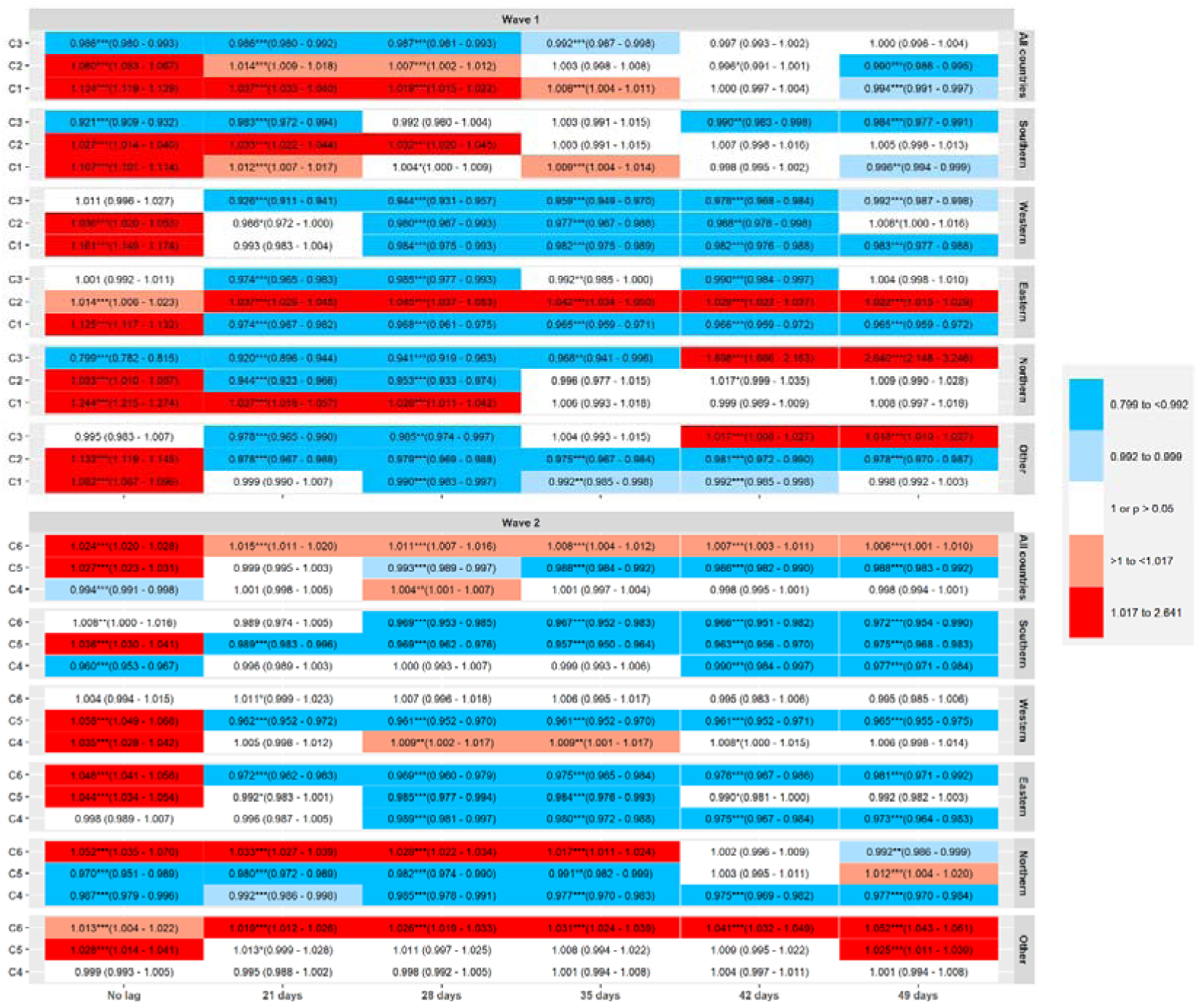
Incidence Rate Ratios (IRR) for the lagged-effects (21, 28, 35, 42 and 49 days) on incident SARS-CoV-2 associated deaths of the three principal component (PCA) scores of NPIs, N= 506795 deaths in 32 countries during the first and second waves of infections (March-December 2020), by region All countries: includes all countries; Southern: Portugal, Spain, Italy, Greece and Cyprus; Western: Belgium, Netherlands, France, Germany, Ireland, United Kingdom and Austria; Eastern: Czech Republic, Slovakia, Slovenia, Poland, Romania, Hungary and Bulgaria; Northern: Norway, Sweden, Finland and Denmark; Other: Croatia, Estonia, Iceland, Latvia, Lithuania, Luxembourg, Malta, Switzerland and Liechtenstein. Within each regional stratum and column for time lags, IRR estimates represent the independent effects of C1-C3 (wave 1) and C4-C6 (wave 2), i.e. mutually adjusted for one another, and additionally adjusted for the immediate effect of each component (no lag), time, 7 day rate change in incidence, and country as random intercept.

In the Southern region during the first wave, a significant (p<0.01) reduction in deaths was only noted for C1 after a 49-days lag, and the same C1 was associated with an increase in the non-lagged variables and after 21 and 35 days. C2 also showed significant associations with an increase in deaths after 21 and 28 days. C3 significantly reduced deaths in the non-lagged model and in models lagged at 21, 42 and 49 days.

In the Western region, C1 showed a significant increase in deaths in non-lagged model and significantly reduced deaths after 28, 35, 42 and 49 days, while C2 significantly reduced deaths after 28, 35 and 42 days. C3 significantly reduced deaths for all time lags considered.

In the Eastern region, C1 increased deaths in the “no-lag” model, but reduced deaths for all the time-lags considered. C2 increased deaths for all time-lags (including the non-lagged model). C3 reduced incident deaths after 21, 28, 35 and 42 days.

In the Northern region, C1 increased deaths in the non-lagged model and after 21 and 28 days. C2 increased deaths in the non-lagged model and significantly reduced deaths after 21 and 28 days. C3 significantly reduced incident deaths in the non-lagged model and after 21, 28 and 35 days lag, but increased deaths after 42 and 49 days (although with large confidence-intervals for the latter two estimates).

In the “other” countries, C1 and C2 showed initially significant associations with increased number of deaths (no-lag). C1 unfolded significant associations with reduced number of deaths after 28, 35 and 42 days, while C2 unfolded significant associations with reduced number of deaths for all the time-lags considered. C3 significantly reduced incident deaths after 21 and 28 days, but turned into significantly associated with an increase in deaths after 42 and 49 days.

During the second wave in all countries (Figure 4), C4 significantly reduced deaths only in the non-lagged model (IRR, 95%CI=0.994, 0.991-0.998), and showed a significant association with increased number of deaths after 28 days (1.004, 1.001-1.007). C5 significantly reduced incident deaths after 28, 35, 42 and 49 days (having started associated with increased deaths in the non-lagged model). C6 showed significant association with increased deaths for all lags considered.

In Southern countries, C4 significantly reduced deaths in the non-lagged model and after 42 and 49 days. C5 increased deaths in the non-lagged model, but significantly reduced deaths for all the time-lags considered. C6 showed an association with increase, and turned into a significant association with a decrease after 28, 35, 42 and 49 days.

In the Western region, C4 showed significant effects with an increase in deaths for all time-lags except after 21, 42 and 49 days. C5 increased deaths in the non-lagged model, but significantly reduced incident deaths for all time-lags considered thereafter, while C6 did not reveal any significant effect on deaths in this groups of countries.

In the Eastern region, C4 significantly reduced deaths following 28, 35, 42 and 49 days, while C5 significantly reduced deaths after 28 and 35 days. C6 significant reduced deaths for all time-lags.

In the Northern region, C4 significantly reduced deaths for all time-lags analysed. C5 significantly reduced deaths for all time-lags except after 42 days (non-significant) and 49 days. C6 showed was associated with an increase in deaths for all time-lags (no-lag, 21, 28 and 35-days) that turned into a significant reduction of deaths after 49 days.

In the group of “Other” countries analysed, C4 did not show any significant effect, while C5 significantly increased deaths in the non-lagged model and after 49 days. C6 showed an association with an increase in deaths for all the time-lags considered.

Supplementary tables S5, S6 show all estimates obtained from models fitted for all countries, tables S10, S11 show these stratified by country groups and tables.

### Sensitivity analysis

The analysis using NPIs collected by OxCGRT showed similar results to those obtained with NPIs collected by COV-PPM for all countries and in the analysis stratified by region, for cases and deaths, Figure 5 and Figure 6 (supplementary Figure S10 and S11 show IRR estimates and 95% CI).

**Fig. 5.**
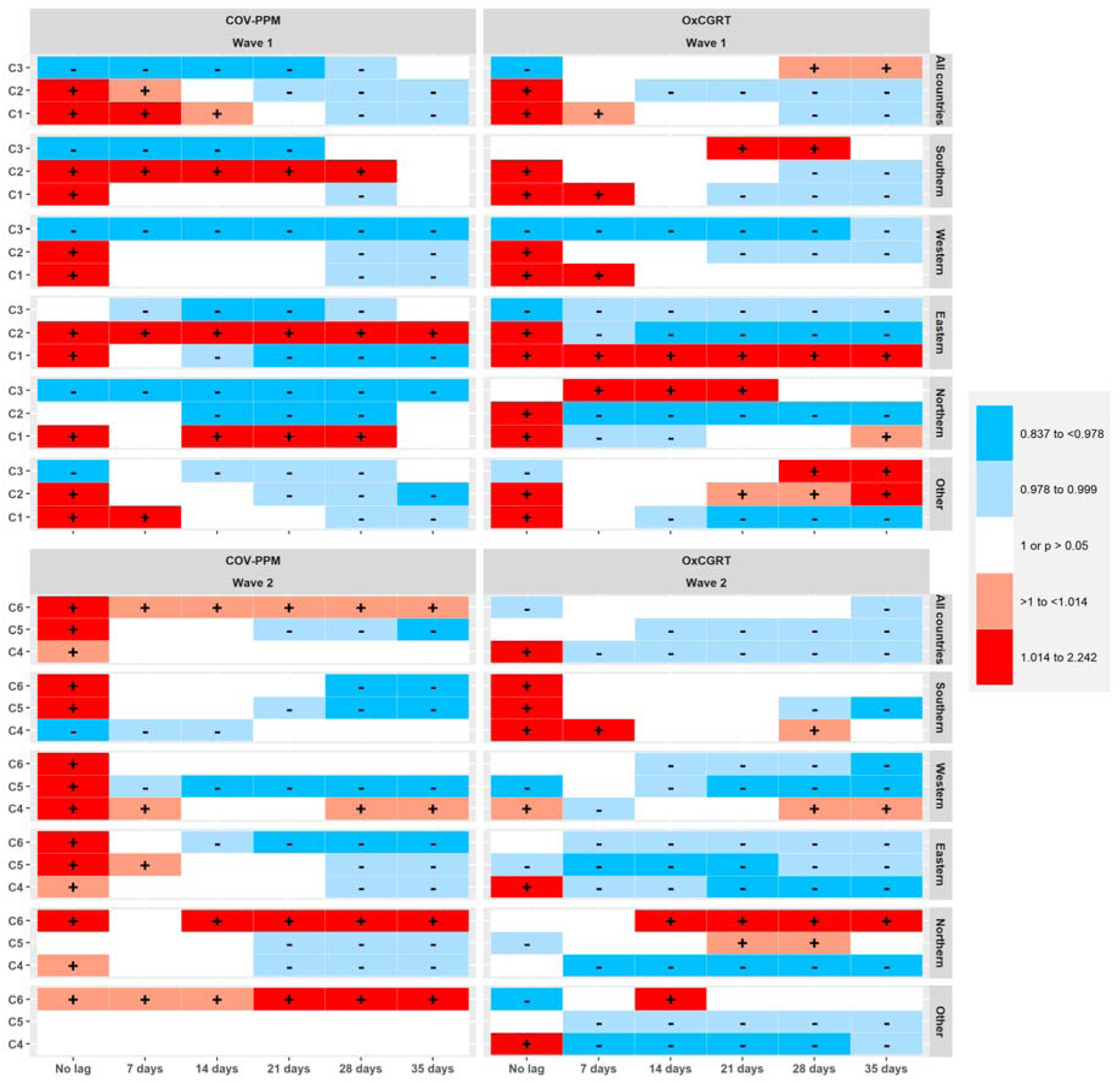
Heat map comparing estimates obtained with COV-PPM and OxCGRT for models predicting incident cases of SARS-Cov-2 (i.e., Table 3 and Table S16 – only colour codes)

**Fig. 6.**
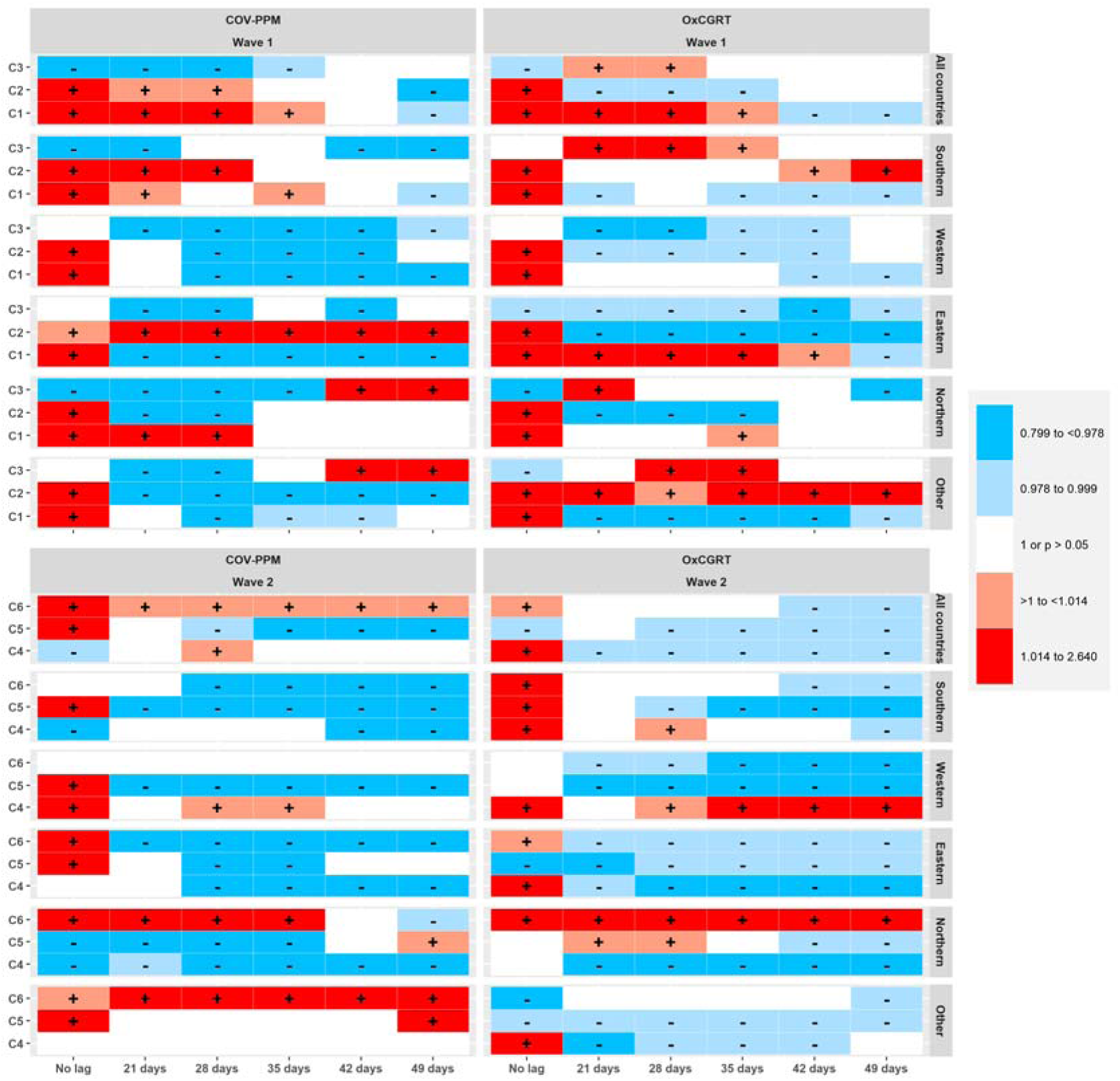
Heat map comparing estimates obtained with COV-PPM and OxCGRT for models predicting deaths of SARS-Cov-2 (i.e. Table 4 and Table S18 – only colour codes)

During the first wave, C1-Ox and C2-Ox, were positively associated with incident cases of SARS-Cov-2 when considering their “non-lagged” effect, and then showed significant negative associations after 21-days, across all countries (supplementary Figure S10). C3-Ox revealed a significant negative association during the first wave for the non-lagged effect, which turned into a significant positive association following 21, 28 and 35 days. During the second wave, C4-Ox revealed a significant positive direct (i.e., non-lagged) effect, that turned into a significant negative effect for all time-lags analysed, again across all countries, and C5-Ox was significantly negatively associated with cases for all time-lags considered. During the second wave, C6-Ox was initially negatively associated with incident cases (no-lag) and also after 35 days, across all countries.

The stratification according to regions obtained using OxCGRT components, in general suggested similar patterns as those obtained with the components summarised using COV-PPM’ group of measures, as shown in Figure 5.

Regarding deaths (supplementary Figure S11) and across all countries, C1-Ox and C2-Ox were positively associated during the first wave considering a “non-lagged” effect, while C3-Ox showed a negative “non-lagged” effect. During this wave, C2-Ox turned to a negative effect following 21, 28 and 35 days, while C1-Ox showed positive associations for 21, 28 and 35 days, that unfolded to significantly negative association after 42 and 49 days. C3-Ox showed significant positive associations after 21, 28 and 35 days.

During the second wave, C4-Ox and C6-Ox showed a positive “non-lagged” effect that became negative after 21 days (C4-Ox) and 35 days (C6-Ox) and remained significant for the following time-lags analysed, across all countries. C5-Ox was negatively associated with deaths across all countries for all the time-lags analysed. Again, the regional stratification showed patterns of associations similar to those obtained with COV-PPM, shown in Figure 6).

## Discussion

In this study we analysed the impact of NPIs, prospectively recorded across 32 European countries, on SARS-Cov-2 incidence and deaths during two pandemic waves of 2020 using principal components derived from two different NPI trackers (COV-PPM and OxCGRT) as exposures. During the first wave of infection, the three component factors (summarizing the effects for all the NPIs analysed) reduced the incidence of SARS-Cov-2 across all countries, albeit with varying time lags. In the second wave, only restrictions to movement/mobility, public transport and healthcare system improvements (C2) were significantly associated with a reduction in incident cases across all countries.

Regional stratification allowed to differentiate the patterns of these impacts for each wave, and showed, for example, that the component C3 related to “masks” was consistently associated with lower incidence of SARS-CoV-2 cases in all regions during the first wave. However, such associations with reduced incidence were only noted in the Southern and Eastern regions during the second wave, while an inverse effect was found in all other regions for at least one of the time-lags considered (adjusted for all other NPI effects).

For deaths, our results support a causal impact of most measures and a time-lagged decrease in mortality. An exception was the effect of C3 (masks) during the second wave, which showed an association with increased number of deaths when considering all countries. Only in Southern and Eastern regions (and Northern after 49 days) could masks significantly reduce deaths, showing again the importance of regional variation in NPI impacts.

We obtained consistent results when analysing SARS-Cov-2 cases and deaths using OxCGRT as exposure, with some exceptions. NPIs contributed to a reduction in cases and deaths, and the exceptions to this pattern observed according to geographical regions, were congruent when comparing both NPIs trackers: for example, during wave 1, COV-PPM’ C2 (healthcare system improvements, border closures) and OxCGRT’ C1-Ox (international travel control, public events, schools, workplace restrictions) did not suggest a reduction in cases in the Eastern region, and also did not suggest a reduction in deaths in the same region (except C1-Ox, but only after 49 days). The same was seen for C6 (masks) and C5-Ox (facial coverings), but also for C6-Ox (public transport measures), which were not associated with a reduction in cases in the Northern region during wave 2. For C3 (masks) and C3-Ox (facial coverings) during wave 1 and across all countries, a reduction in cases was initially observed, which disappeared after 35 days using COV-PPM data and which turned positive after 28 and 35 days with OxCGRT, and a very close pattern was noticed for deaths during this wave. However, during the second wave, across all countries, only C5-Ox (facial coverings), did suggest a reduction in cases and deaths.

Overall, the negative impact of NPIs on SARS-Cov-2 incidence (i.e., the associations with a reduction in incidence, expressed as IRR below the unit) suggests that the pandemic control strategies were effective in reducing, at least partially, the incidence across these countries at population-level. It should be noted that our observations of “what worked” may only be valid for the observed periods in 2020, i.e., where the population was immunologically naive, had no access to vaccination and was faced with a respiratory virus with a comparable natural course of diseases or similar reproduction rate. With the emergence and roll-out of COVID-19 vaccines, the inference of our effectiveness estimates beyond this time period may be limited. However, in case of escape variants that evade immunity acquired through vaccines or previous infections, the knowledge generated from this study could inform the design and combination of future NPIs to protect immunologically naive populations against the threat of respiratory virus.

Our results concur with and add to previous observations of the effect of physical distance interventions imposed across 149 countries, which were associated with a reduction in COVID-19 incidence.[4]

The recommended or compulsory use of masks seemed to consistently reduce SARS-Cov-2 cases and deaths at the population-level according to our findings, with such effect during the first wave of infection being noticeable for all the time-lagged effects considered and across all countries. This is in line with a review showing the potential effectiveness of mask utilization in community settings, particularly in the case of specific mask types (i.e. medical types),[15] with a recent review on the effectiveness of NPIs to reduce SARS-Cov-2 transmission[16] and with a cluster-randomized trial conducted in Bangladesh that showed the community-level effect of masks distribution on SARS-Cov-2 cases.[17] The fact that “masks” were not consistently associated with a reduction in the number of SARS-CoV-2 cases during the second wave across and all countries may be due to the homogeneity of this measure during this period, with almost all countries (except Northern countries) recommending the use of facial masks in that time period so that variance was substantially reduced and proper counterfactuals were lacking in the second wave to detect the effect of the measure.

The stratified analysis by waves also revealed different magnitudes of effects for the same components, i.e. the same set of combinations of NPIs, included when comparing waves. These differences may be due to organizational safety measures and individual protective behaviours in place already during the first wave, thus reducing the additional effect of newer implementations, as previously suggested [18]. Again, this can also help explain the seemingly strong effect observed for recommended or mandatory use of “masks” in the Northern region during the first wave, reflecting the late and looser approach taken to NPIs implementation in some of the countries in this region.

Other measures, such as border closures or travelling restrictions, as part of other NPIs, seemed effective particularly in the first wave, but not in all regions. Their effectiveness may strongly depend on the timing of implementation, which is also in line with the results of a Cochrane review that included mainly modelling studies suggesting that such measures may lead to a reduction in the number of new cases, although with large uncertainty and when implemented at the beginning of the outbreak[19]. Travel restrictions were also analysed using the ECDC categorization of NPIs for the European region[1], showing different impacts, dependent on the starting date and on the combination of NPIs in which they have been implemented[20] (e.g., quarantine of incoming travellers, enforcement of hygiene concepts and limitations to and from specified high-risk regions).

Across all waves and regions, the analysed NPI components also showed non-significant effects or significant associations with higher incidences or deaths. These non-significant effects may suggest that the respective measures had no measurable effect at the population level, when adjusted for the immediate effect and the co-occurrence of other NPI combinations. However, this does not allow the inference that a given measure is completely ineffective at a smaller scale (e.g. in a given region or town of a country) or at the individual-level. Although we adjusted for the 7-day preceding change rate in incidence, reverse causation for the immediate (non-lagged) effects that show associations with higher incidences and deaths cannot be completely ruled out as it is more likely that measures were taken *because* of high infections rates or deaths, and not that measures *caused* higher infections and deaths. In the lagged models, positive associations (i.e. IRR above the unit, or associated with an increase) on incidence or deaths mean that the measures were not able (e.g. due to improper design, implementation, stringency, or adherence) to change pre-existing rising trends of infections or deaths. The results from models fitted only with the lagged effects on incidence across all countries (without the immediate effect variables, results not shown), support our interpretation by showing the same pattern of associations. However, we cannot rule out residual confounding by other unmeasured variables that may well lie in the causal chain explaining the effect of NPIs on the outcomes.

Nevertheless, the use of two different exposures (despite also the uncertainty about the delay between NPIs implementation), constitutes an attempt to reduce the potential bias stemming from the use of different outcomes (e.g. incomplete cases, different testing capacities), which has been rarely tested in other studies [2].

Further explorations, e.g. through structural equation models, are needed to more clearly elucidate the causal pathways related to NPIs and pandemic control, such as the role of media communication, political discourses, individual behaviour, social support networks, or trust.

### Strengths and Limitations

We used a standardized procedure to monitor and code NPIs taken across Europe and resorted to a natural experiment approach[21], which stands as a suitable and strong research design to monitor and analyse the effects of the measures to control the pandemic. Implemented in a panel design, each country serves as its own control while cross-national differences are considered simultaneously. Our estimates are, however, limited by the fact that they do not disentangle the effect of each NPI *individually*, but assessed the independent effect of three sets of NPI *combinations* that explained a high proportion of the variance across Europe. In other words, the use of principal component analysis to summarize data means that we can only refer to the effect of the group of related measures in each component, limiting the attribution of direct effects to specific categories of NPIs. Despite this limitation, a principal component analysis is regarded *state of the art* to address the challenge of multiple co-occurring policies in effectiveness studies[22]. Future studies should analyse the independent effects of individual NPIs, and also explore the use of other outcome measures such as the reproduction number.

We further cannot rule-out the possibility of misclassification of NPIs, particularly within some of the subcategories of the measures established. Furthermore, we did not account for the heterogeneity in the baseline status of, for e.g., healthcare infrastructure, that could determine a larger or smaller effect of any particular measure to improve the healthcare system response. However, the congruency in the effects of some of the NPIs summarized in the stratified analysis by regions, is in favour of an independent effect across different constellations of systems, despite the needed (and legitimate) simplification by attributing the same meaning to each group of NPIs for each of the 32 countries.

Other structured data collection efforts are being developed by different consortia, categorizing NPIs across the globe using different criteria and aggregation levels and updated with different periodicity[23–26], allowing to further assess NPIs effectiveness. We conducted a sensitivity analysis with selected NPIs covered in the OxCGRT[10], and obtained patterns that confirm our findings, despite the inevitable differences in NPIs categorisations.

The use of notified cases and deaths might be a further limitation of this study, due potential changes in national notification processes or testing intensity, for example, which can directly impact these outcomes. However, the use of different lag periods to measure the impact of NPIs on the different outcomes (including the analysis with “no-lags” provided as supplement), and the congruency in the results obtained using the two different exposures, can also be seen as a sensitivity analysis to the potential bias imposed by outcome data quality, as previously suggested by authors calling for rigorous impact analysis in this field[3]. Nonetheless, further testing should be done in subsequent studies, to rule-out any impact of changes in surveillance system, case ascertainment, or cause of death attribution rules, for example.

An analysis conducted with the “Stringency index” of NPIs proposed in the scope of the OxCGRT, showed that the timing of restrictive measures implementations seems crucial to mitigate SARS-Cov-2 incidence[27], but strategies did not have the same effect in all the countries with available data. Another analysis relating the “Stringency Index” aggregated at the continental level, with COVID-19 Case-fatality Rates (CFR) worldwide, did not observe a statistically significant association between the Index and COVID-19 CFR[28]. The authors found that stricter measures were associated with higher CFR in high-income countries with active testing policies (testing anyone symptomatic or testing open to public), suggesting that more restrictive (lockdown) measures might hit the most vulnerable groups harder[28]. This calls for further research that considers socioeconomic (and inequality) aspects of the measures taken for pandemic control.

As NPIs are implemented and withdrawn dynamically, attempts to track these interventions need to embrace a continuous effort within an appropriate monitoring framework.[18] A modelling effort conducted for 16 different countries, suggest that implementation of dynamic interventions (i.e., alternating between periods of NPIs enforcement followed by periods of relaxation), might not be ideal[29]. In another analysis, the combination of physical distancing measures implemented with varying intensity and timing (border controls, restriction on mass gatherings, lockdown type measures) seem to be effective if implemented early (about two weeks before the 100^th^ case), although individual effect is hard to disentangle since several measures were implemented very close to one another[30].

Changes in NPIs effectiveness across time are expected since their stringency changes (e.g. from recommendations to impositions with envisaged punishment for noncompliance), measures are refined (e.g., continuous adaptation of infrastructures to ensure social distance, new air ventilators/purifiers) and the population also adapts to and evolves in the way measures are understood and followed. The effectiveness may hence be very time- and context-dependent. This adds to the complexity in the understanding of NPIs’ effectiveness, and substantiates the research challenges ahead to provide relevant information for public health decision-making.

## Statements and Declarations

### Authors contribution

Conceptualisation: DC, SR, KB

Data curation: DC, SR, KB

Formal analysis: DC

Writing of first and final draft: DC, KB

Revision for important intellectual content: SR, KB

### Ethics approval and consent to participate

The information gathered and analysed was retrieved from public official websites across Europe and from anonymised publicly available datasets, namely SARS-Cov-2 infection cases and deaths, retrieved from the public World Health Organization online platform. No ethical clearance was required.

### Consent to publish

Not applicable.

### Availability of data statement

The NPIs database can be provided by the corresponding author upon reasonable request.

## Supporting information

Additonal file 1

## Data Availability

The information gathered and analysed was retrieved from public official websites across Europe and from freely available datasets. The NPIs database used can be accessed at https://doi.org/10.7910/DVN/ANTOH7.
SARS-Cov-2 infection cases and deaths was retrieved from the public WHO online platform: https://covid19.who.int/data.

## Acknowledgements

The authors would like to acknowledge all researchers (Amir Mohsenpour, Andreas Gold, Victoria Saint, Rosa Jahn, Niklas Nutsch, Stella Duwendag) and student assistants (Anna Mundry, Catharina Mühling, Johanna Nonte, Annalena Sander, Helena Haskia, Leonie Klose, Nicole Krzisczyk, Oxana Klassen, Julia Winkelnkemper, Cathrin Stöber, Neelab Hakim, Sarah Markgraf, Sukhvir Kaur, Lilian Salis, Theresa Herzig, Kübra Altinok, Anne Krämer) who helped collecting and correcting the data about the NPIs being implemented across Europe. We also acknowledge the comments of colleagues from the Robert Koch Institute, Matthias an der Heiden, Andreas Hicketier and Viviane Bremer, on earlier versions of this manuscript.

