## Supplementary material for "Impact of non-pharmaceutical interventions on COVID-19 incidence and deaths: cross-national natural experiment in 32 European countries": Additonal file 1

**Index**

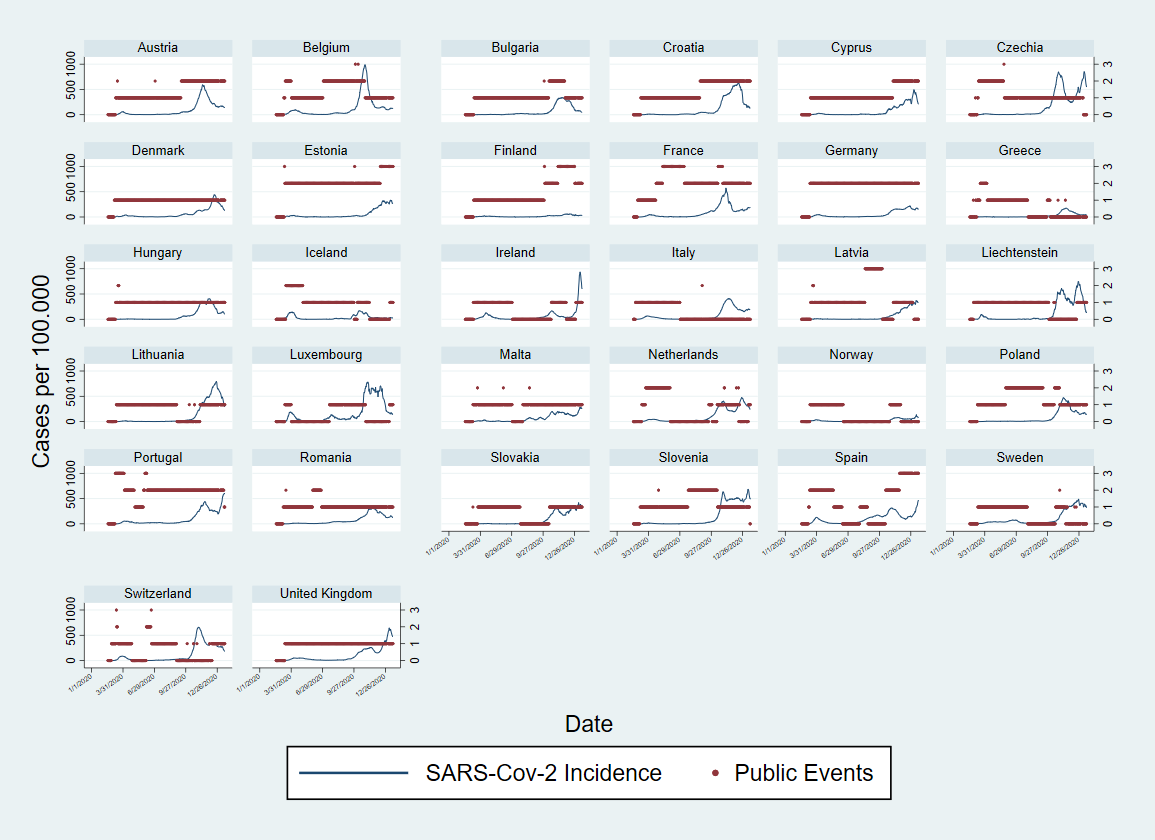

### Figure S1. Plots showing categories of measures related to “Public Events” in place during the observation period (March-December 2020) and SARS-Cov-2 incidence per 100,000, in 32 countries from the European region.

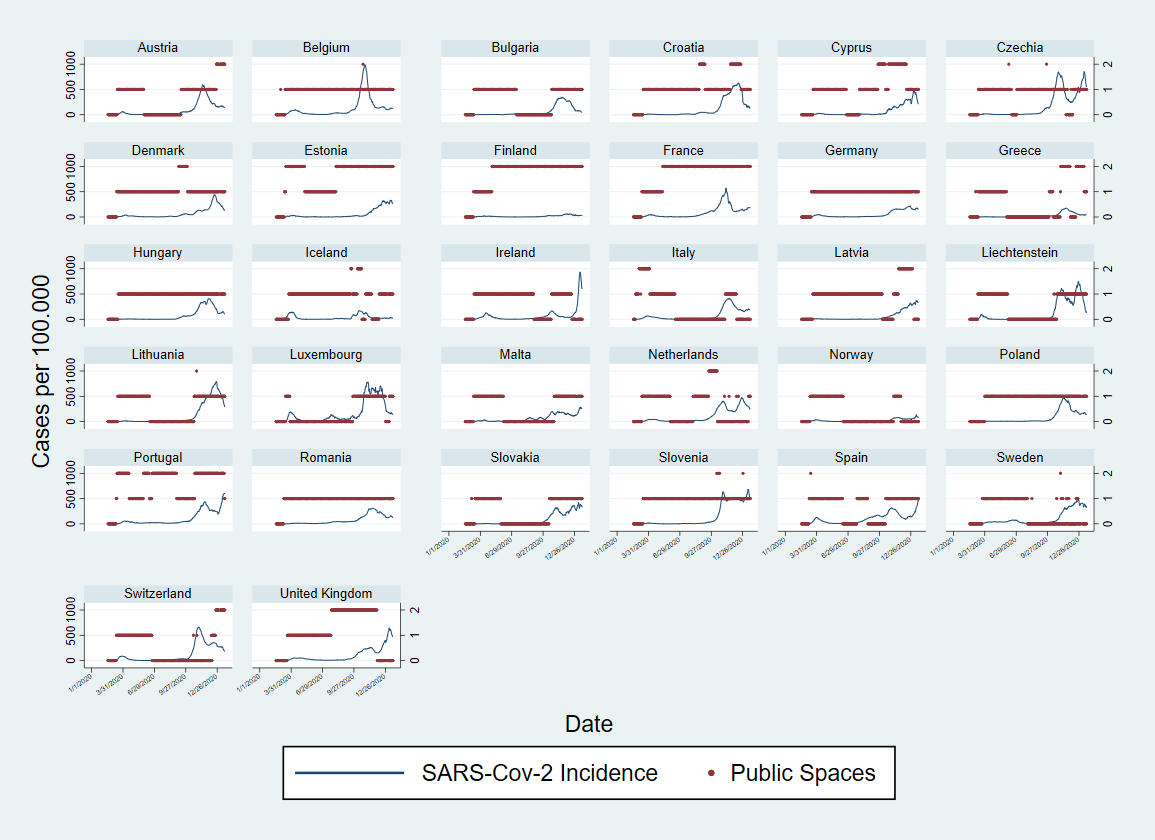

### Figure S2. Plots showing categories of measures related to “Public Spaces” in place during the observation period (March-December 2020) and SARS-Cov-2 incidence per 100,000, in 32 countries from the European region.

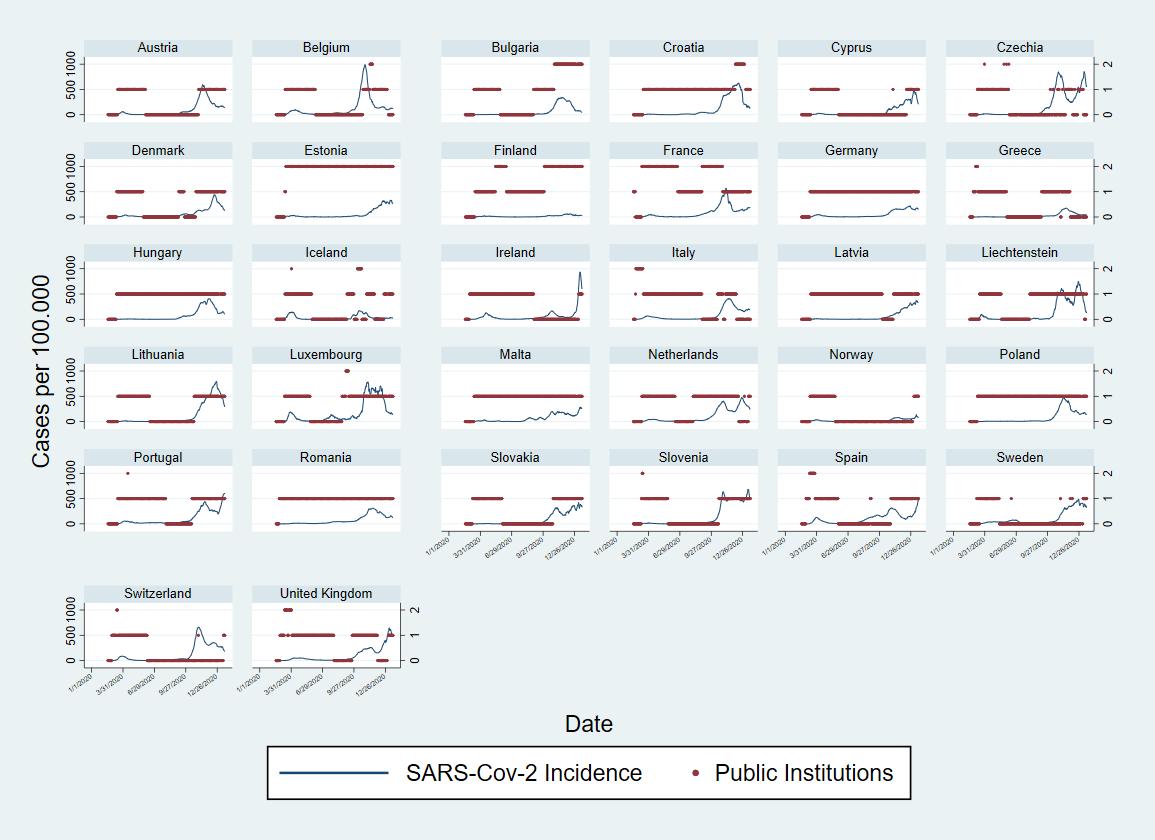

### Figure S3. Plots showing categories of measures related to “Public Institutions” in place during the observation period (March-December 2020) and SARS-Cov-2 incidence per 100,000, in 32 countries from the European region.

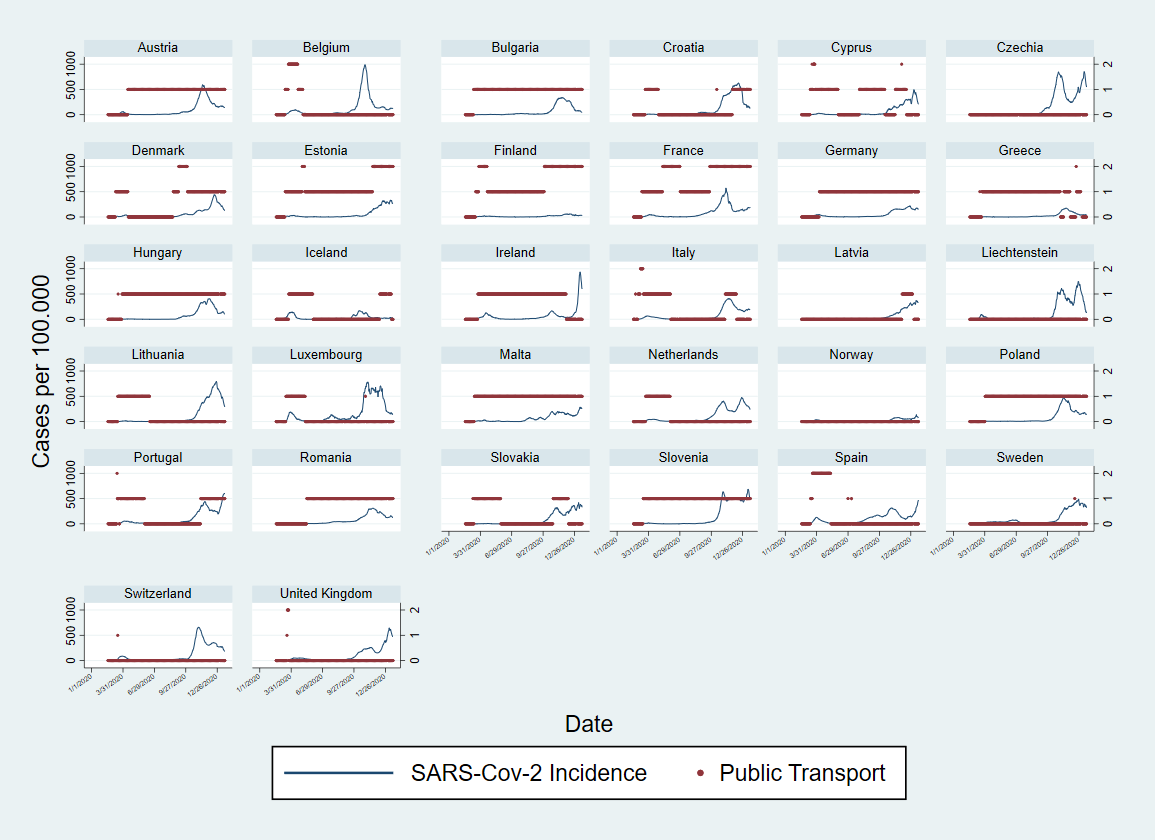

### Figure S4. Plots showing categories of measures related to “Public transport” in place during the observation period (March-December 2020) and SARS-Cov-2 incidence per 100,000, in 32 countries from the European region.

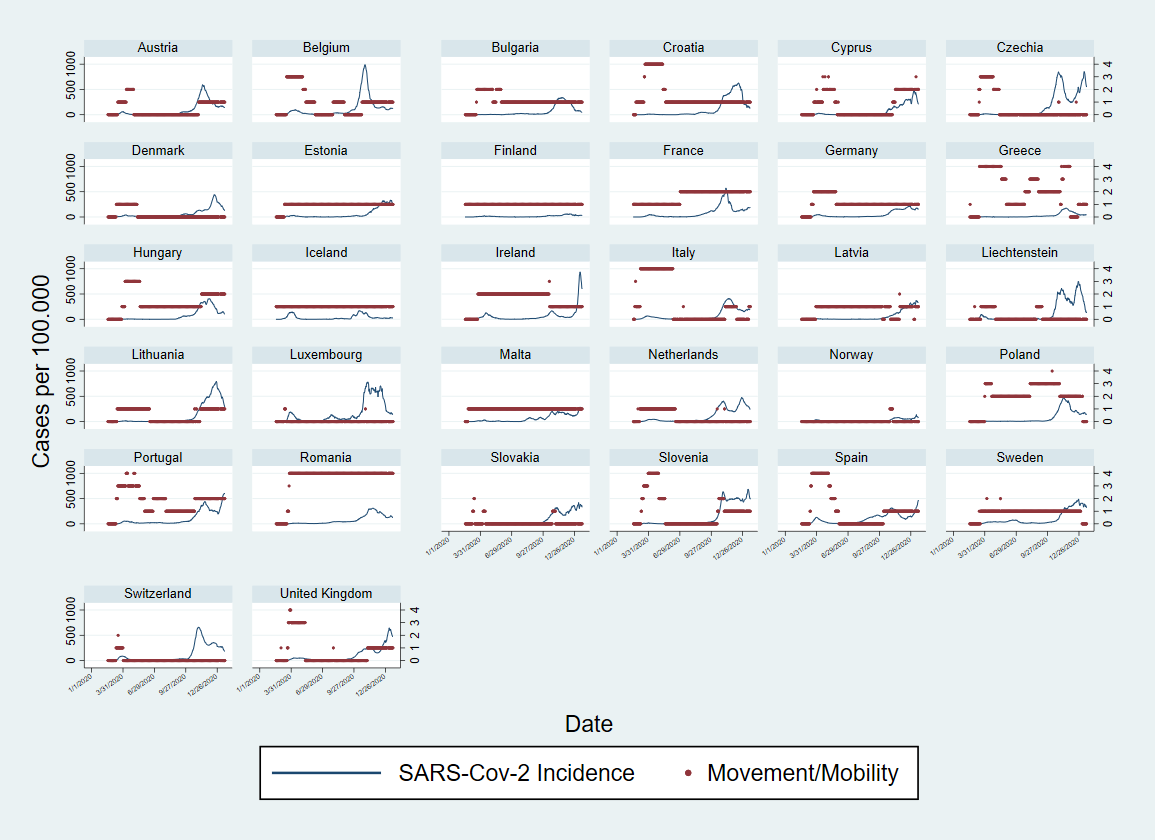

### Figure S5. Plots showing categories of measures related to “Movement/mobility” in place during the observation period (March-December 2020) and SARS-Cov-2 incidence per 100,000, in 32 countries from the European region.

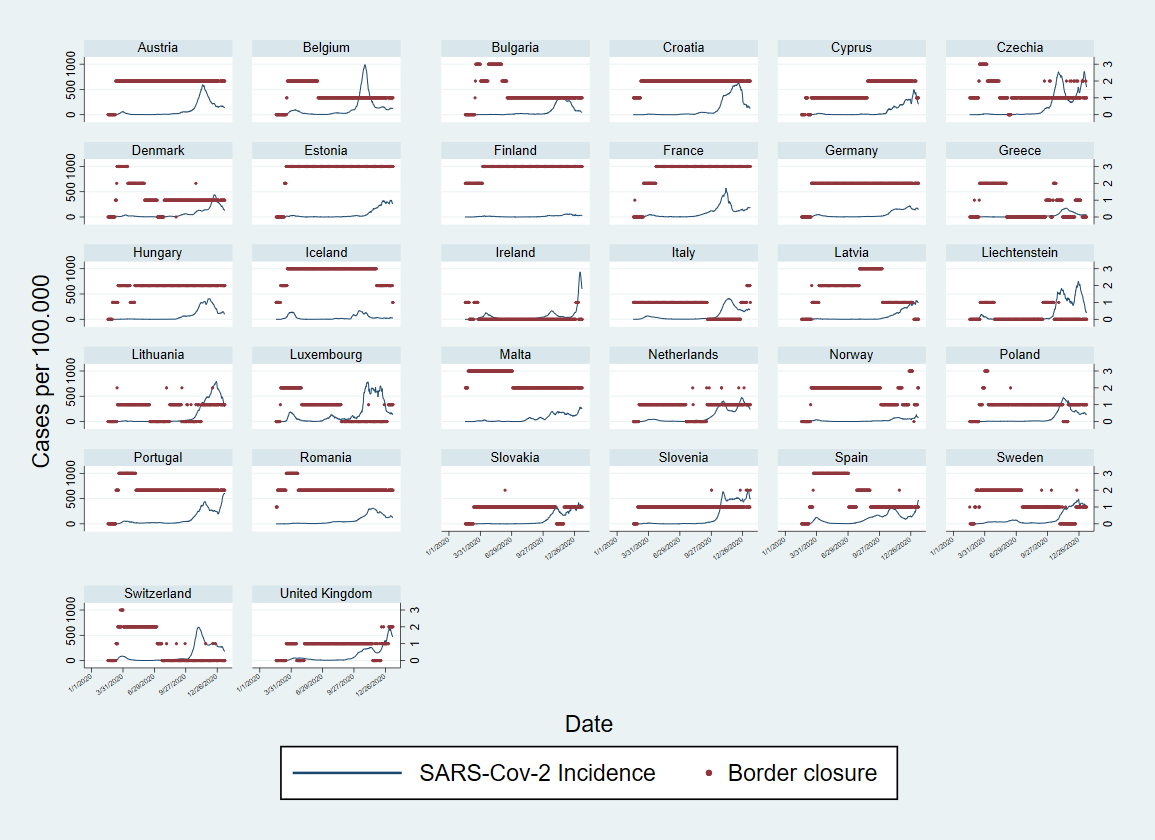
Figure S6. Plots showing categories of measures related to “Border closure” in place during the observation period (March-December 2020) and SARS-Cov-2 incidence per 100,000, in 32 countries from the European region.

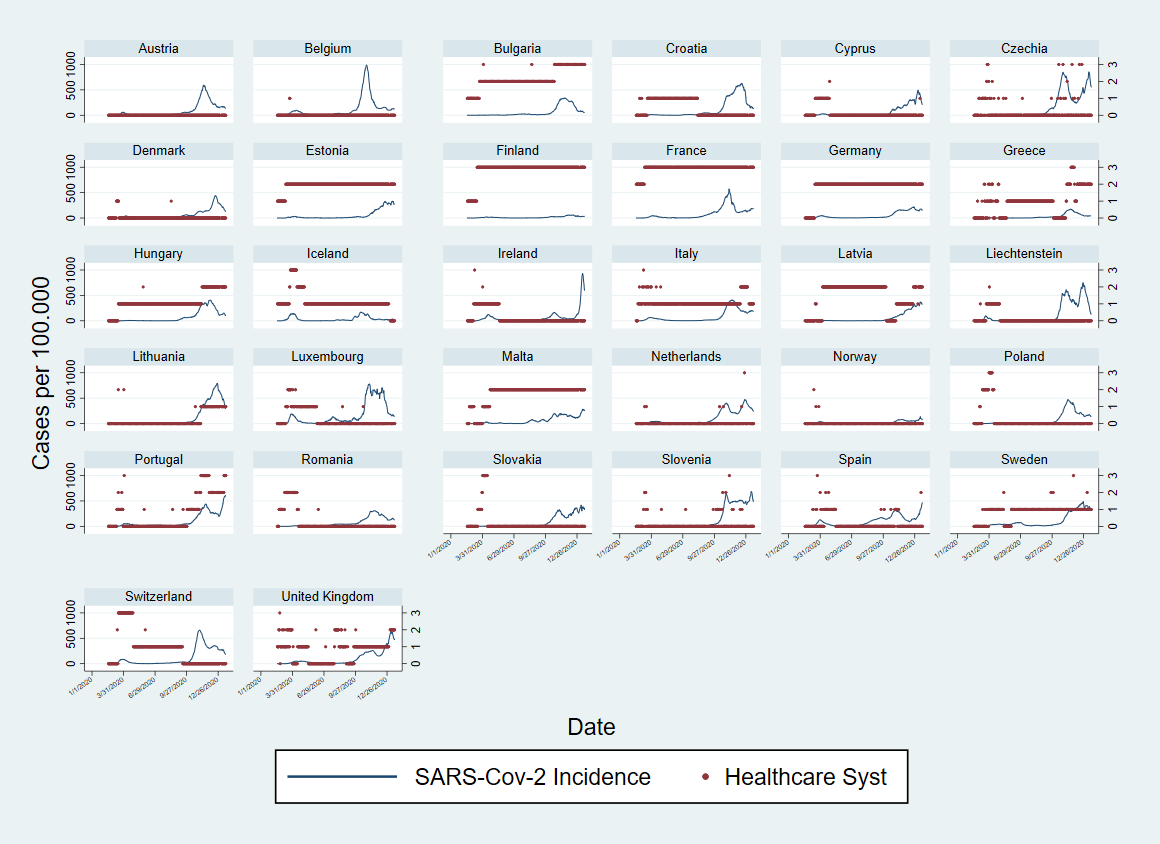
Figure S7. Plots showing categories of measures related to “Improvement to the healthcare system” in place during the observation period (March-December 2020) and SARS-Cov-2 incidence per 100,000, in 32 countries from the European region.

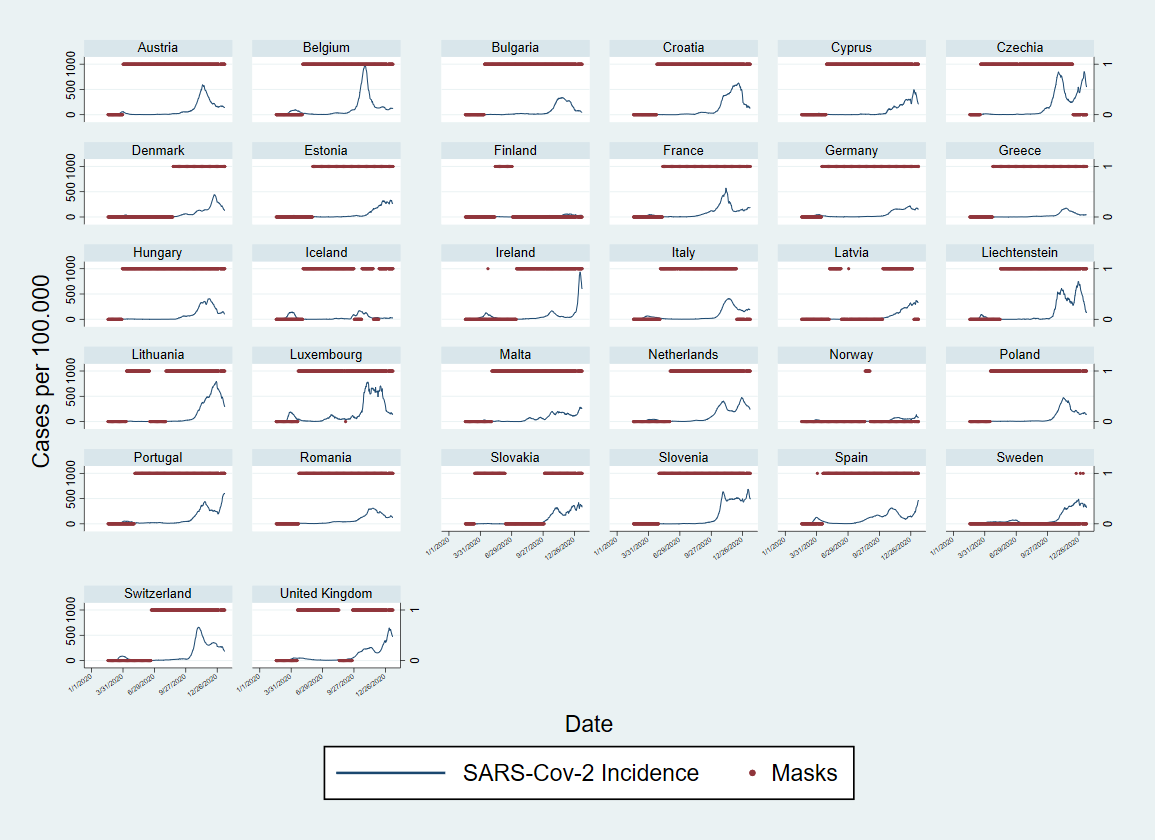

### Figure S8. Plots showing measures aiming at “Masks” recommendation/enforcement during the observation period (March-December 2020) and SARS-Cov-2 incidence per 100.000, in 32 countries from the European region.

| Wave 1 | Wave 2 |
| --- | --- |
| 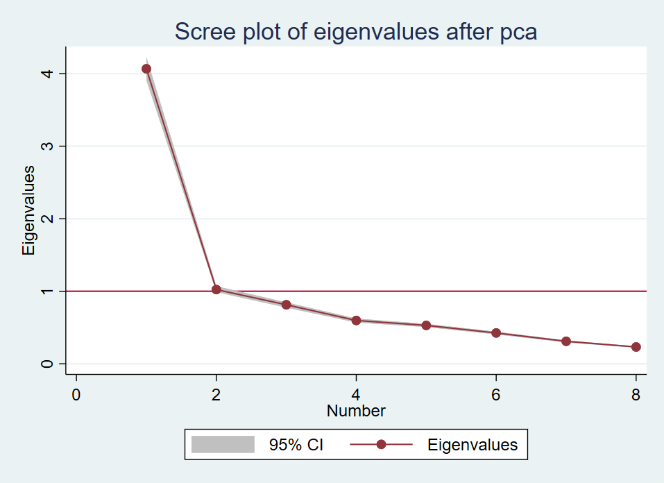 | 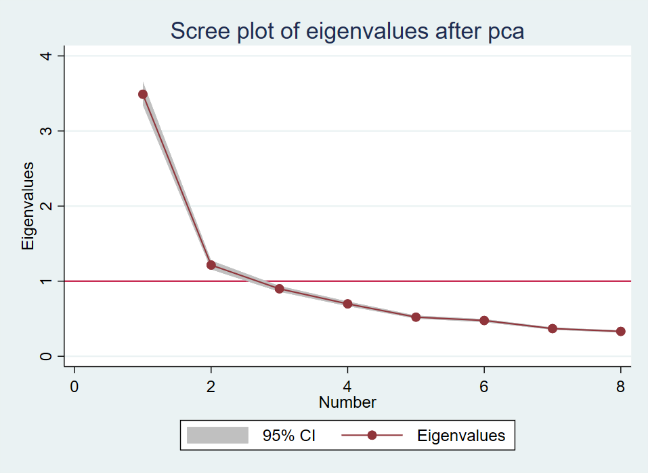 |

### Figure S9. Scree plots from PCA conducted over selected non-pharmaceutical interventions (NPIs) for Wave 1 (calendar weeks 5-25, February to June) and Wave 2 (calendar weeks 35-52 end of August to December) during 2020.

### Panel S1. Categorization of non-pharmaceutical interventions variables for analysis

**a) Public events:** coded as 0 if no measure was in place; coded as 1 if at least one of the categories “not-specified”, “more than 1000 persons”, “less than 1000 persons”, “More than 50 persons”, “any type/number” was present; coded as 2 if at least two of these categories were simultaneously present; coded as 3 if three or four of these categories (unless not mutually exclusive) were simultaneously present.

**b) Public institutions:** coded as 0 if no measure was in place; coded as 1 if at least one of the categories “not-specified”, “single cities”, “state level” or “national”, was present; coded as 2 if two or more these categories were simultaneously present;

**c) Public spaces:** coded as 0 if no measure was in place; coded as 1 if at least one of the categories “not-specified”, “single cities”, “state level” or “national”, was present; coded as 2 if two or more these categories were simultaneously present;

**d) Public transports:** coded as 0 if no measure was in place; coded as 1 if at least one of the categories “not-specified”, “single cities”, “state level” or “national”, was present; coded as 2 if two or more these categories were simultaneously present;

**e) Movement/mobility:** coded as 0 if no measure was in place; coded as 1 if at least one of the categories “not specified”, “pedestrians”, “private cars”, “aviation national travel”, or “other (Ships, trains, etc)”, was present; coded as 2 if two of these categories were simultaneously present; coded as 3 if three of these categories were simultaneously present; code as 4 if four or more of these categories were simultaneously present;

**f) Travelling or border closure:** coded as 0 if no measure was in place; coded as 1 if at least one of the categories “not specified”, “for non-nationals from high-risk regions”, “for all non-nationals”, or “for all incoming travellers” was present; coded as 2 if two of these categories were simultaneously present; coded as 3 if three of more of these categories were simultaneously present;

**g) Measures to improve the healthcare system:** coded as 0 if no measure was in place; coded as 1 if at least one of the categories “not specified”, “HR reinforcement or redistribution”, “technical reinforcement or redistribution”, “material infrastructural reinforcement or redistribution” was present; coded as 2 if two of these categories were simultaneously present; coded as 3 if three or more of these categories were simultaneously present.

**h) Masks:** coded as 0 if no measure was in place; coded as 1 if any measure for nose and mouth protection recommendation or mandatory requirement, was in place.

### Table S1. Variation observed in the eight non-pharmaceutical interventions (NPIs) used, categories described in Panel S1, and the three component scores, across the 32 countries analysed, by wave.

| Wave 1 |  |  |  |  |  |  |  | Wave 2 |  |  |  |  |
| --- | --- | --- | --- | --- | --- | --- | --- | --- | --- | --- | --- | --- |
| Variable |  | Mean | Std. Dev. | Min | Max | Observations | | Mean | Std. Dev. | Min | Max | Observations |
| Public events | overall | 0.8471514 | 0.7085490 | 0 | 3 | N = 4704 | | 1.077538 | 0.819334 | 0 | 3 | N = 4256 |
|  | between |  | 0.2894207 | 0.1292517 | 1.47619 | n = 32 |  |  | 0.666409 | 0.015038 | 2.180451 | n = 32 |
|  | within |  | 0.6487509 | -0.6290391 | 3.19409 | T = 147 |  |  | 0.4909 | -0.4563 | 3.0625 | T = 133 |
| Public spaces | overall | 0.6940901 | 0.5845112 | 0 | 2 |  |  | 0.7434211 | 0.6192782 | 0 | 2 |  |
|  | between |  | 0.2315859 | 0.4013605 | 1.489796 |  |  |  | 0.4840963 | 0.0075188 | 2 |  |
|  | within |  | 0.5382251 | -0.7957058 | 2.156675 |  |  |  | 0.3955077 | -0.7077068 | 2.156955 |  |
| Public institutions | overall | 0.6753827 | 0.5627363 | 0 | 2 |  |  | 0.9309211 | 0.6937171 | 0 | 2 |  |
|  | between |  | 0.1853622 | 0.0884354 | 1.061224 |  |  |  | 0.5169163 | 0.0827068 | 2 |  |
|  | within |  | 0.5323342 | -0.3858418 | 2.022321 |  |  |  | 0.471519 | -0.5878759 | 2.848214 |  |
| Public transport | overall | 0.4491922 | 0.5669802 | 0 | 2 |  |  | 0.5596805 | 0.6110693 | 0 | 2 |  |
|  | between |  | 0.2805871 | 0 | 0.9795918 |  |  |  | 0.5497136 | 0 | 1.774436 |  |
|  | within |  | 0.495158 | -0.5303997 | 2.374362 |  |  |  | 0.2838928 | -0.2222744 | 1.93562 |  |
| Movement/mobility | overall | 1.017857 | 1.235634 | 0 | 4 |  |  | 0.8858083 | 0.9433424 | 0 | 4 |  |
|  | between |  | 0.673215 | 0 | 2.687075 |  |  |  | 0.8560943 | 0 | 4 |  |
|  | within |  | 1.042901 | -1.669218 | 3.963435 |  |  |  | 0.4239501 | -1.06156 | 2.93844 |  |
| Border closure | overall | 1.344175 | 1.044592 | 0 | 3 |  |  | 1.345395 | 0.9350556 | 0 | 3 |  |
|  | between |  | 0.6281309 | 0.2040816 | 2.802721 |  |  |  | 0.8853125 | 0.0300752 | 3 |  |
|  | within |  | 0.8419451 | -0.7034439 | 3.487032 |  |  |  | 0.3389178 | -0.2786654 | 2.841635 |  |
| Healthcare system improvement | overall | 0.7055697 | 0.9230682 | 0 | 3 |  |  | 0.7882989 | 1.012989 | 0 | 3 |  |
|  | between |  | 0.6744655 | 0 | 2.693878 |  |  |  | 0.951732 | 0 | 3 |  |
|  | within |  | 0.6413058 | -0.9270833 | 3.542304 |  |  |  | 0.3852965 | -0.6101974 | 3.743186 |  |
| Masks | overall | 0.329932 | 0.4702381 | 0 | 1 |  |  | 0.8601974 | 0.3468229 | 0 | 1 |  |
|  | between |  | 0.1832463 | 0 | 0.6462585 |  |  |  | 0.2920201 | 0 | 1 |  |
|  | within |  | 0.4342662 | -0.3163265 | 1.323129 |  |  |  | 0.1940511 | -0.1322838 | 1.84516 |  |
| Component 1 (4 for wave 2) | overall | 50 | 16.96749 | 24.32622 | 95.29994 |  |  | 50 | 16.62913 | 21.62831 | 93.31548 |  |
|  | between |  | 6.786782 | 35.64553 | 63.18238 |  |  |  | 15.1712 | 27.99734 | 84.95831 |  |
|  | within |  | 15.59696 | 14.25135 | 86.26042 |  |  |  | 7.314572 | 21.897 | 72.01978 |  |
| Component 2 (5 for wave 2) | overall | 50 | 14.1044 | 30.54177 | 92.62379 |  |  | 50 | 12.7371 | 29.41809 | 86.60291 |  |
|  | between |  | 8.569364 | 40.56193 | 75.04967 |  |  |  | 11.75188 | 34.91665 | 77.89664 |  |
|  | within |  | 11.30397 | 15.0046 | 84.77342 |  |  |  | 5.330221 | 26.34875 | 73.42411 |  |
| Component 3 (6 for wave 2) | overall | 50 | 10.62755 | 36.12392 | 72.27434 |  |  | 50 | 11.01743 | 16.23185 | 62.31834 |  |
|  | between |  | 3.703322 | 41.66169 | 57.89436 |  |  |  | 9.61353 | 20.07576 | 60.33374 |  |
|  | within |  | 9.982789 | 32.7591 | 76.43093 |  |  |  | 5.64189 | 16.20047 | 75.6832 |  |

### Table S2. Test of model specifications

First, models were adjusted for a variable of day-count and the natural logarithm of the population size as offset. Second, models were tested with the three component scores and finally with the 7-day incidence rate change variable. Negative binomial, Poisson and gamma distributions were tested, considering country as a random or fixed effects factor in the case of negative binomial link.

Wave 1

| Model (observations, n=4480, weeks 5-25, 2020) | Family | LL(model) | df | AIC | BIC |
| --- | --- | --- | --- | --- | --- |
| Cases_ms7~Day + offset \|\| (country as random) | nbinomial | -30326.06 | 4 | 60660.11 | 60685.94 |
| Cases_ms7~Day + offset + Country (as fixed) | nbinomial | -30251.74 | 34 | 60571.47 | 60790.98 |
| Cases_ms7~Day + offset + Country (as fixed) | poisson | -6731096 | 33 | 1.35e+07 | 1.35e+07 |
| Cases_ms7~Day + offset + Country (as fixed) | gamma | -1.78e+14 | 33 | 3.55e+14 | 3.55e+14 |
| Cases_ms7~Day + C1+C2+C3 + offset \|\| (country as random) | nbinomial | -28611.36 | 7 | 57236.72 | 57281.91 |
| Cases_ms7~Day + C1+C2+C3 + offset + Country (as fixed) | nbinomial | -28519.36 | 37 | 57112.72 | 57351.60 |
| Cases_ms7~Day + C1+C2+C3 + offset + Country (as fixed) | poisson | -3361023 | 36 | 6722118 | 6722350 |
| Cases_ms7~Day + C1+C2+C3 + offset + Country (as fixed) | gamma | -28441.62 | 36 | 56955.23 | 57187.66 |
| Cases_ms7~Day + C1+C2+C3 + rate_change + offset \|\| (country as random) | nbinomial | -28479.72 | 8 | 56975.44 | 57027.09 |
| Cases_ms7~Day + C1+C2+C3 + rate_change + offset + Country (as fixed) | nbinomial | -28386.80 | 38 | 56849.59 | 57094.93 |
| Cases_ms7~Day + C1+C2+C3 + rate_change + offset + Country (as fixed) | poisson | -3165739 | 37 | 6331552 | 6331791 |
| Cases_ms7~Day + C1+C2+C3 + rate_change + offset + Country (as fixed) | gamma | -28029.40 | 37 | 56132.79 | 56371.67 |

Wave 2

| Model (observations, n=032 weeks 35-52, 2020) | Family | LL(model) | df | AIC | BIC |
| --- | --- | --- | --- | --- | --- |
| Cases_ms7~Day + offset \|\| (country as random) | nbinomial | -41313.40 | 4 | 82634.79 | 82660.22 |
| Cases_ms7~Day + offset + Country (as fixed) | nbinomial | -41221.70 | 34 | 82511.40 | 82727.50 |
| Cases_ms7~Day + offset + Country (as fixed) | poisson | -2.15e+07 | 33 | 4.29e+07 | 4.29e+07 |
| Cases_ms7~Day + offset + Country (as fixed) | gamma | -1.17e+15 | 33 | 2.35e+15 | 2.35e+15 |
| Cases_ms7~Day + C4+C5+C6 + offset \|\| (country as random) | nbinomial | -41154.49 | 7 | 82322.98 | 82367.47 |
| Cases_ms7~Day + C4+C5+C6 + offset + Country (as fixed) | nbinomial | -41059.32 | 37 | 82192.64 | 82427.81 |
| Cases_ms7~Day + C4+C5+C6 + offset + Country (as fixed) | poisson | -1.95e+07 | 36 | 3.90e+07 | 3.90e+07 |
| Cases_ms7~Day + C4+C5+C6 + offset + Country (as fixed) | gamma | -41802.66 | 36 | 83677.33 | 83906.15 |
| Cases_ms7~Day + C4+C5+C6 + rate_change + offset \|\| (country as random) | nbinomial | -41016.29 | 8 | 82048.59 | 82099.44 |
| Cases_ms7~Day + C4+C5+C6 + rate_change + offset + Country (as fixed) | nbinomial | -40910.82 | 38 | 81915.65 | 82157.18 |
| Cases_ms7~Day + C4+C5+C6 + rate_change + offset + Country (as fixed) | poisson | -1.90e+07 | 37 | 3.81e+07 | 3.81e+07 |
| Cases_ms7~Day + C4+C5+C6 + rate_change + offset + Country (as fixed) | gamma | -41746.21 | 37 | 83566.42 | 83801.59 |

### Table S3. Incidence Rate Ratios (IRR) – cases - for the lagged-effects obtained for the three extracted components from PCA of NPIs (pc1 – pc3), considering a 7, 14, 21, 28 and 35 days-lag across all countries – Wave 1 (weeks 5-25 or end of February-June, 2020).

| **Cases** - Wave 1 (32 countries) |  |  |  |  |  |  |
| --- | --- | --- | --- | --- | --- | --- |
| VARIABLES | IRR (95%CI) | IRR (95%CI) | IRR (95%CI) | IRR (95%CI) | IRR (95%CI) | IRR (95%CI) |
| ID_day | 1.027***(1.025 - 1.029) | 1.026***(1.023 - 1.028) | 1.022***(1.020 - 1.025) | 1.018***(1.015 - 1.020) | 1.012***(1.009 - 1.014) | 1.003**(1.001 - 1.005) |
| Tpc1nolagw1 | 1.111***(1.106 - 1.115) | 1.090***(1.082 - 1.098) | 1.093***(1.088 - 1.099) | 1.092***(1.087 - 1.097) | 1.086***(1.082 - 1.091) | 1.073***(1.069 - 1.077) |
| Tpc2nolagw1 | 1.051***(1.045 - 1.057) | 1.041***(1.034 - 1.049) | 1.045***(1.039 - 1.051) | 1.042***(1.037 - 1.048) | 1.034***(1.029 - 1.039) | 1.026***(1.021 - 1.031) |
| Tpc3nolagw1 | 0.962***(0.955 - 0.969) | 0.985***(0.976 - 0.995) | 0.982***(0.974 - 0.990) | 0.981***(0.974 - 0.988) | 0.983***(0.977 - 0.989) | 0.987***(0.982 - 0.993) |
| Tpc1_7lagw1 |  | 1.017***(1.010 - 1.024) |  |  |  |  |
| Tpc2_7lagw1 |  | 1.011***(1.003 - 1.018) |  |  |  |  |
| Tpc3_7lagw1 |  | 0.968***(0.959 - 0.978) |  |  |  |  |
| Tpc1_14lagw1 |  |  | 1.008***(1.002 - 1.013) |  |  |  |
| Tpc2_14lagw1 |  |  | 1.002(0.996 - 1.008) |  |  |  |
| Tpc3_14lagw1 |  |  | 0.968***(0.961 - 0.976) |  |  |  |
| Tpc1_21lagw1 |  |  |  | 1.002(0.997 - 1.006) |  |  |
| Tpc2_21lagw1 |  |  |  | 0.993***(0.987 - 0.998) |  |  |
| Tpc3_21lagw1 |  |  |  | 0.975***(0.968 - 0.981) |  |  |
| Tpc1_28lagw1 |  |  |  |  | 0.995**(0.992 - 0.999) |  |
| Tpc2_28lagw1 |  |  |  |  | 0.987***(0.982 - 0.992) |  |
| Tpc3_28lagw1 |  |  |  |  | 0.988***(0.982 - 0.994) |  |
| Tpc1_35lagw1 |  |  |  |  |  | 0.994***(0.990 - 0.997) |
| Tpc2_35lagw1 |  |  |  |  |  | 0.984***(0.979 - 0.988) |
| Tpc3_35lagw1 |  |  |  |  |  | 1.001(0.996 - 1.007) |
| ratechange | 1.100***(1.084 - 1.117) | 1.100***(1.083 - 1.117) | 1.066***(1.051 - 1.081) | 1.031***(1.021 - 1.042) | 1.005(0.998 - 1.013) | 0.987***(0.982 - 0.992) |
| lnalpha | 1.726***(1.656 - 1.800) | 1.643***(1.576 - 1.713) | 1.548***(1.484 - 1.615) | 1.398***(1.340 - 1.459) | 1.177***(1.127 - 1.228) | 0.891***(0.853 - 0.931) |
| var(_cons[Country_code]) | 4.707***(2.189 - 10.125) | 4.521***(2.144 - 9.536) | 4.166***(2.056 - 8.439) | 3.766***(1.953 - 7.262) | 3.389***(1.851 - 6.202) | 3.068***(1.761 - 5.343) |
| Observations | 4.704 | 4.576 | 4.352 | 4.128 | 3.904 | 3.680 |
| 95% CI – Confidence Intervals in parentheses |  |  |  |  |  |  |
| *** p<0.01, ** p<0.05, * p<0.1 |  |  |  |  |  |  |

### Table S4. Incidence Rate Ratios (IRR) – cases - for the lagged-effects obtained for the three extracted components from PCA of NPIs (pc4 – pc6), considering a 7, 14, 21, 28 and 35 days-lag across all countries – Wave 2 (weeks 35-52 or end of August-December, 2020).

| **Cases** - Wave 2 (32 countries) |  |  |  |  |  |  |
| --- | --- | --- | --- | --- | --- | --- |
| VARIABLES | IRR (95%CI) | IRR (95%CI) | IRR (95%CI) | IRR (95%CI) | IRR (95%CI) | IRR (95%CI) |
| ID_day | 1.025***(1.024 - 1.025) | 1.024***(1.024 - 1.025) | 1.025***(1.024 - 1.025) | 1.025***(1.024 - 1.026) | 1.025***(1.025 - 1.026) | 1.026***(1.025 - 1.026) |
| Tpc4nolagw2 | 1.004**(1.001 - 1.007) | 1.004**(1.001 - 1.008) | 1.004***(1.001 - 1.008) | 1.004**(1.001 - 1.007) | 1.004**(1.001 - 1.007) | 1.004***(1.001 - 1.007) |
| Tpc5nolagw2 | 1.022***(1.018 - 1.026) | 1.019***(1.014 - 1.024) | 1.022***(1.018 - 1.026) | 1.023***(1.019 - 1.027) | 1.025***(1.021 - 1.028) | 1.023***(1.020 - 1.027) |
| Tpc6nolagw2 | 1.023***(1.019 - 1.026) | 1.015***(1.009 - 1.020) | 1.018***(1.014 - 1.022) | 1.018***(1.014 - 1.022) | 1.017***(1.013 - 1.021) | 1.018***(1.014 - 1.021) |
| Tpc4_7lagw2 |  | 1.000(0.996 - 1.003) |  |  |  |  |
| Tpc5_7lagw2 |  | 1.003(0.998 - 1.008) |  |  |  |  |
| Tpc6_7lagw2 |  | 1.012***(1.007 - 1.017) |  |  |  |  |
| Tpc4_14lagw2 |  |  | 0.998(0.995 - 1.002) |  |  |  |
| Tpc5_14lagw2 |  |  | 0.997(0.993 - 1.002) |  |  |  |
| Tpc6_14lagw2 |  |  | 1.010***(1.006 - 1.014) |  |  |  |
| Tpc4_21lagw2 |  |  |  | 0.999(0.996 - 1.002) |  |  |
| Tpc5_21lagw2 |  |  |  | 0.990***(0.986 - 0.994) |  |  |
| Tpc6_21lagw2 |  |  |  | 1.011***(1.007 - 1.015) |  |  |
| Tpc4_28lagw2 |  |  |  |  | 1.000(0.997 - 1.003) |  |
| Tpc5_28lagw2 |  |  |  |  | 0.980***(0.976 - 0.984) |  |
| Tpc6_28lagw2 |  |  |  |  | 1.011***(1.007 - 1.014) |  |
| Tpc4_35lagw2 |  |  |  |  |  | 0.999(0.996 - 1.002) |
| Tpc5_35lagw2 |  |  |  |  |  | 0.975***(0.971 - 0.979) |
| Tpc6_35lagw2 |  |  |  |  |  | 1.008***(1.005 - 1.012) |
| ratechange | 1.494***(1.421 - 1.571) | 1.508***(1.434 - 1.586) | 1.492***(1.418 - 1.569) | 1.475***(1.403 - 1.551) | 1.466***(1.395 - 1.540) | 1.474***(1.403 - 1.548) |
| lnalpha | 0.389***(0.374 - 0.406) | 0.387***(0.372 - 0.403) | 0.387***(0.372 - 0.403) | 0.386***(0.370 - 0.402) | 0.381***(0.365 - 0.396) | 0.377***(0.362 - 0.393) |
| var(_cons[Country_code]) | 1.545***(1.241 - 1.923) | 1.542***(1.239 - 1.919) | 1.476***(1.210 - 1.799) | 1.374***(1.168 - 1.616) | 1.296***(1.136 - 1.479) | 1.267***(1.125 - 1.427) |
| Observations | 4.256 | 4.256 | 4.256 | 4.256 | 4.256 | 4.256 |
| 95% CI – Confidence Intervals in parentheses |  |  |  |  |  |  |
| *** p<0.01, ** p<0.05, * p<0.1 |  |  |  |  |  |  |

### Table S5. Incidence Rate Ratios (IRR) -deaths- for the lagged-effects obtained for the three extracted components from PCA of NPIs (pc1 – pc3), considering a 21, 28, 35, 42 and 49 days-lag across all countries – Wave 1 (weeks 5-25 or end of February-June, 2020).

| **Deaths** - Wave 1 (32 countries) | |  | |  | |  | |  | |  | |
| --- | --- | --- | --- | --- | --- | --- | --- | --- | --- | --- | --- |
| VARIABLES | IRR (95%CI) | | IRR (95%CI) | | IRR (95%CI) | | IRR (95%CI) | | IRR (95%CI) | | IRR (95%CI) |
| ID_day | 1.027***(1.025 - 1.029) | | 1.011***(1.008 - 1.013) | | 1.010***(1.007 - 1.013) | | 1.007***(1.004 - 1.010) | | 0.997*(0.995 - 1.000) | | 0.992***(0.989 - 0.995) |
| Tpc1nolagw1 | 1.124***(1.119 - 1.129) | | 1.086***(1.081 - 1.090) | | 1.092***(1.088 - 1.097) | | 1.088***(1.083 - 1.092) | | 1.064***(1.059 - 1.069) | | 1.042***(1.038 - 1.046) |
| Tpc2nolagw1 | 1.060***(1.053 - 1.067) | | 1.053***(1.046 - 1.059) | | 1.048***(1.042 - 1.055) | | 1.034***(1.028 - 1.041) | | 1.015***(1.009 - 1.021) | | 0.996(0.990 - 1.002) |
| Tpc3nolagw1 | 0.986***(0.980 - 0.993) | | 0.987***(0.981 - 0.993) | | 0.989***(0.983 - 0.995) | | 0.992**(0.986 - 0.999) | | 0.993**(0.988 - 0.999) | | 0.994**(0.989 - 0.999) |
| Tpc1_21lagw1 |  | | 1.037***(1.033 - 1.040) | |  | |  | |  | |  |
| Tpc2_21lagw1 |  | | 1.014***(1.009 - 1.018) | |  | |  | |  | |  |
| Tpc3_21lagw1 |  | | 0.986***(0.980 - 0.992) | |  | |  | |  | |  |
| Tpc1_28lagw1 |  | |  | | 1.019***(1.015 - 1.022) | |  | |  | |  |
| Tpc2_28lagw1 |  | |  | | 1.007***(1.002 - 1.012) | |  | |  | |  |
| Tpc3_28lagw1 |  | |  | | 0.987***(0.981 - 0.993) | |  | |  | |  |
| Tpc1_35lagw1 |  | |  | |  | | 1.008***(1.004 - 1.011) | |  | |  |
| Tpc2_35lagw1 |  | |  | |  | | 1.003(0.998 - 1.008) | |  | |  |
| Tpc3_35lagw1 |  | |  | |  | | 0.992***(0.987 - 0.998) | |  | |  |
| Tpc1_42lagw1 |  | |  | |  | |  | | 1.000(0.997 - 1.004) | |  |
| Tpc2_42lagw1 |  | |  | |  | |  | | 0.996*(0.991 - 1.001) | |  |
| Tpc3_42lagw1 |  | |  | |  | |  | | 0.997(0.993 - 1.002) | |  |
| Tpc1_49lagw1 |  | |  | |  | |  | |  | | 0.994***(0.991 - 0.997) |
| Tpc2_49lagw1 |  | |  | |  | |  | |  | | 0.990***(0.986 - 0.995) |
| Tpc3_49lagw1 |  | |  | |  | |  | |  | | 1.000(0.996 - 1.004) |
| ratechange | 0.952***(0.942 - 0.962) | | 0.953***(0.944 - 0.962) | | 0.929***(0.920 - 0.939) | | 0.905***(0.895 - 0.914) | | 0.761***(0.744 - 0.779) | | 0.707***(0.688 - 0.727) |
| lnalpha | 1.219***(1.157 - 1.284) | | 0.932**(0.882 - 0.984) | | 0.985(0.933 - 1.040) | | 0.928***(0.879 - 0.981) | | 0.689***(0.649 - 0.732) | | 0.533***(0.500 - 0.568) |
| var(_cons[Country_code]) | 13.632***(3.751 - 49.537) | | 12.643***(3.612 - 44.255) | | 9.808***(3.173 - 30.315) | | 7.622***(2.794 - 20.796) | | 5.411***(2.351 - 12.455) | | 4.665***(2.181 - 9.979) |
| Observations | 4.704 | | 4.128 | | 3.904 | | 3.680 | | 3.456 | | 3.232 |
| 95% CI – Confidence Intervals in parentheses |  | |  | |  | |  | |  | |  |
| *** p<0.01, ** p<0.05, * p<0.1 |  | |  | |  | |  | |  | |  |

### Table S6. Incidence Rate Ratios (IRR) -deaths- for the lagged-effects obtained for the three extracted components from PCA of NPIs (pc4 – pc6), considering a 21, 28, 35, 42 and 49 days-lag across all countries – Wave 2 (weeks 35-52 or end of August-December, 2020).

| **Deaths** - Wave 2 (32 countries) |  |  |  |  |  |  |
| --- | --- | --- | --- | --- | --- | --- |
| VARIABLES | IRR (95%CI) | IRR (95%CI) | IRR (95%CI) | IRR (95%CI) | IRR (95%CI) | IRR (95%CI) |
| ID_day | 1.034***(1.033 - 1.035) | 1.033***(1.033 - 1.034) | 1.034***(1.033 - 1.034) | 1.034***(1.034 - 1.035) | 1.035***(1.034 - 1.035) | 1.034***(1.034 - 1.035) |
| Tpc4nolagw2 | 0.994***(0.991 - 0.998) | 0.994***(0.991 - 0.997) | 0.995***(0.991 - 0.998) | 0.995***(0.992 - 0.998) | 0.995***(0.992 - 0.998) | 0.994***(0.991 - 0.997) |
| Tpc5nolagw2 | 1.027***(1.023 - 1.031) | 1.027***(1.023 - 1.031) | 1.028***(1.024 - 1.032) | 1.028***(1.024 - 1.032) | 1.027***(1.023 - 1.031) | 1.026***(1.022 - 1.030) |
| Tpc6nolagw2 | 1.024***(1.020 - 1.028) | 1.019***(1.015 - 1.024) | 1.021***(1.017 - 1.025) | 1.020***(1.016 - 1.025) | 1.021***(1.017 - 1.025) | 1.023***(1.019 - 1.027) |
| Tpc4_7lagw2 |  | 1.001(0.998 - 1.005) |  |  |  |  |
| Tpc5_7lagw2 |  | 0.999(0.995 - 1.003) |  |  |  |  |
| Tpc6_7lagw2 |  | 1.015***(1.011 - 1.020) |  |  |  |  |
| Tpc4_14lagw2 |  |  | 1.004**(1.001 - 1.007) |  |  |  |
| Tpc5_14lagw2 |  |  | 0.993***(0.989 - 0.997) |  |  |  |
| Tpc6_14lagw2 |  |  | 1.011***(1.007 - 1.016) |  |  |  |
| Tpc4_21lagw2 |  |  |  | 1.001(0.997 - 1.004) |  |  |
| Tpc5_21lagw2 |  |  |  | 0.988***(0.984 - 0.992) |  |  |
| Tpc6_21lagw2 |  |  |  | 1.008***(1.004 - 1.012) |  |  |
| Tpc4_28lagw2 |  |  |  |  | 0.998(0.995 - 1.001) |  |
| Tpc5_28lagw2 |  |  |  |  | 0.986***(0.982 - 0.990) |  |
| Tpc6_28lagw2 |  |  |  |  | 1.007***(1.003 - 1.011) |  |
| Tpc4_35lagw2 |  |  |  |  |  | 0.998(0.994 - 1.001) |
| Tpc5_35lagw2 |  |  |  |  |  | 0.988***(0.983 - 0.992) |
| Tpc6_35lagw2 |  |  |  |  |  | 1.006***(1.001 - 1.010) |
| ratechange | 0.650***(0.615 - 0.687) | 0.662***(0.626 - 0.700) | 0.651***(0.616 - 0.688) | 0.646***(0.612 - 0.683) | 0.649***(0.615 - 0.686) | 0.651***(0.616 - 0.688) |
| lnalpha | 0.328***(0.312 - 0.344) | 0.323***(0.308 - 0.339) | 0.324***(0.308 - 0.340) | 0.323***(0.307 - 0.339) | 0.322***(0.306 - 0.338) | 0.323***(0.308 - 0.339) |
| var(_cons[Country_code]) | 2.257***(1.509 - 3.377) | 2.106***(1.456 - 3.047) | 2.104***(1.455 - 3.044) | 2.082***(1.448 - 2.995) | 2.082***(1.448 - 2.992) | 2.079***(1.447 - 2.987) |
| Observations | 4.256 | 4.256 | 4.256 | 4.256 | 4.256 | 4.256 |
| 95% CI – Confidence Intervals in parentheses |  |  |  |  |  |  |
| *** p<0.01, ** p<0.05, * p<0.1 |  |  |  |  |  |  |

### Figure S10. Incidence Rate Ratios (IRR) and 95% CI for the lagged-effects (7, 14, 21, 28 and 35 days) on SARS-CoV2 incidence of the three principal component (PCA) scores of NPIs (using OxCGRT data) in 32 countries during the first and second waves of infections (March-December 2020), by region

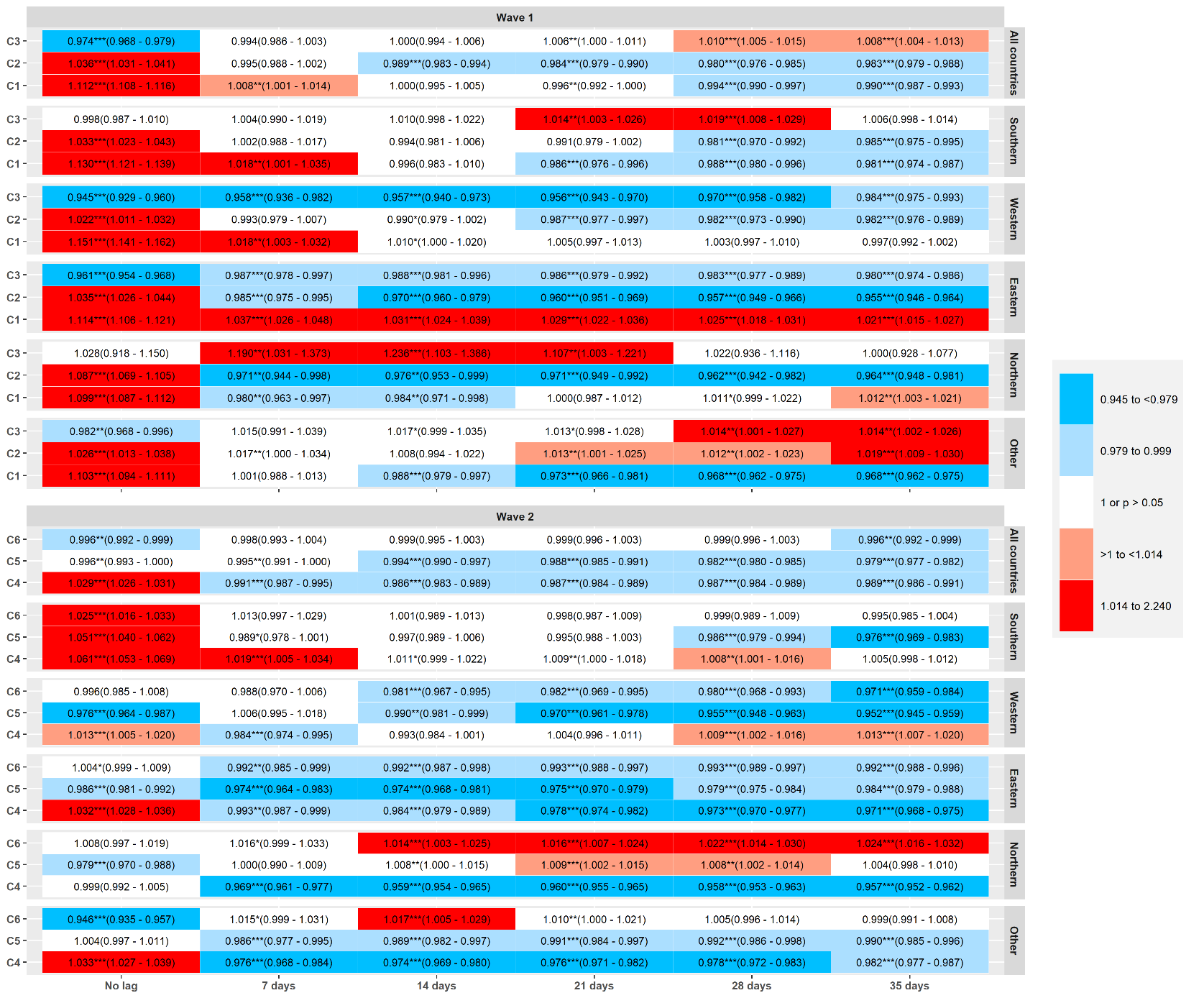

All countries: includes all countries; Southern: Portugal, Spain, Italy, Greece and Cyprus; Western: Belgium, Netherlands, France, Germany, Ireland, United Kingdom and Austria; Eastern: Czech Republic, Slovakia, Slovenia, Poland, Romania, Hungary and Bulgaria; Northern: Norway, Sweden, Finland and Denmark; Other: Croatia, Estonia, Iceland, Latvia, Lithuania, Luxembourg, Malta, Switzerland and Liechtenstein.

### Figure S11. Incidence Rate Ratios (IRR) and 95% CI for the lagged-effects (21, 28, 35, 42 and 49 days) on SARS-CoV2 deaths of the three principal component (PCA) scores of NPIs (using OxCGRT data) in 32 countries during the first and second waves of infections (March-December 2020), by region

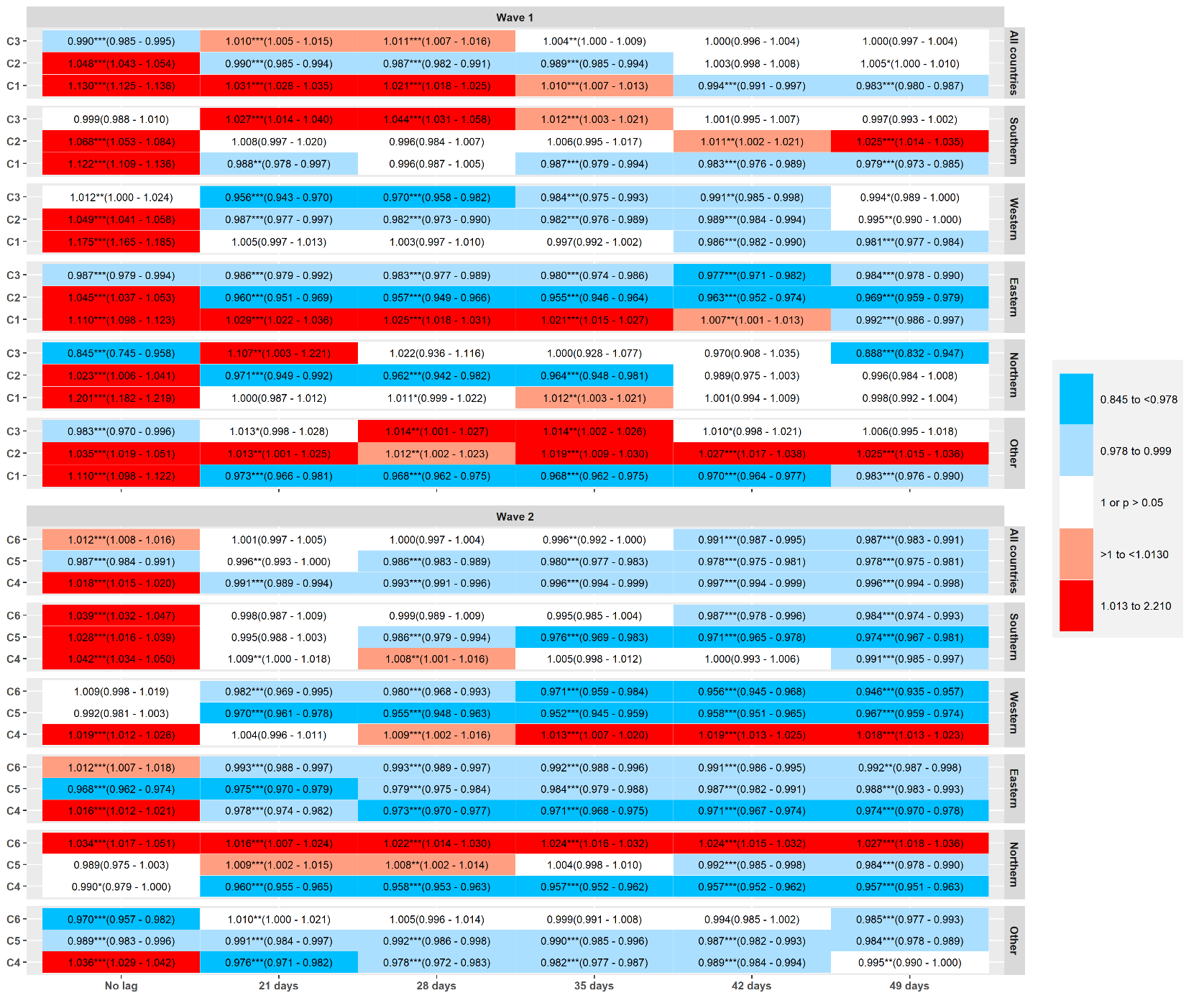

*** p<0.01, ** p<0.05, * p<0.1

All countries: includes all countries; Southern: Portugal, Spain, Italy, Greece and Cyprus; Western: Belgium, Netherlands, France, Germany, Ireland, United Kingdom and Austria; Eastern: Czech Republic, Slovakia, Slovenia, Poland, Romania, Hungary and Bulgaria; Northern: Norway, Sweden, Finland and Denmark; Other: Croatia, Estonia, Iceland, Latvia, Lithuania, Luxembourg, Malta, Switzerland and Liechtenstein.
